# DRAGON-Data: A platform and protocol for integrating genomic and phenotypic data across large psychiatric cohorts

**DOI:** 10.1101/2022.01.18.22269463

**Authors:** Leon Hubbard, Amy J. Lynham, Sarah Knott, Jack F. G. Underwood, Richard Anney, Jonathan I. Bisson, Marianne.B.M van den Bree, Nick Craddock, Michael O’Donovan, Ian Jones, George Kirov, Kate Langley, Joanna Martin, Frances Rice, Neil Roberts, Anita Thapar, Michael J. Owen, Jeremy Hall, Antonio F. Pardiñas, James T.R. Walters

## Abstract

**Introduction:** Current psychiatric diagnoses, although heritable, have not been clearly mapped onto distinct underlying pathogenic processes. The same symptoms often occur in multiple disorders, and a substantial proportion of both genetic and environmental risk factors are shared across disorders. However, the relationship between shared symptomatology and shared genetic liability is still poorly understood. Well-characterised, cross-disorder samples are needed to investigate this matter, but currently few exist, and severe mental disorders are poorly represented in existing biobanking efforts. Purposely curated and aggregated data from individual research groups can fulfil this unmet need, resulting in rich resources for psychiatric research.

**Methods and analyses:** As part of the Cardiff MRC Mental Health Data Pathfinder, we have curated and harmonised phenotypic and genetic information from 15 studies within the MRC Centre for Neuropsychiatric Genetics and Genomics to create a new data repository, DRAGON-DATA. To date, DRAGON-DATA includes over 45,000 individuals: adults or children with psychiatric diagnoses, affected probands with family members and individuals who carry a known neurodevelopmental copy number variant (ND-CNV). We have processed the available phenotype information to derive core variables that can be reliably analysed across groups. In addition, all datasets with genotype information have undergone rigorous quality control, imputation, CNV calling and polygenic score generation.

**Ethics and Dissemination:** DRAGON-DATA combines genetic and non-genetic information and is available as a resource for research across traditional psychiatric diagnostic categories. Its structure and governance follow standard UK ethical requirements (at the level of participating studies and the project as a whole) and conforms to principles reflected in the EU data protection scheme (GDPR). Algorithms and pipelines used for data harmonisation are currently publicly available for the scientific community, and an appropriate data sharing protocol will be developed as part of ongoing projects (DATAMIND) in partnership with HDR UK.

## Introduction

The value of collaboration and data sharing is well recognised within the medical community and is one of the hallmarks of what has been called “the fourth age of research”, in which the pace of discovery has accelerated and international platforms for studying multifactorial problems have been built^12^. The aggregation of data from individual research groups not only maximises the utility of individual datasets and minimises demands on participants, but enables the joint analyses of complex data that can lead to incremental advances in elucidating disease aetiology^3^. Within major psychiatric and neurodevelopmental conditions, few truly novel pharmacological treatments have been developed for several decades, with the noteworthy exceptions of ketamine for depression^4^ and atomoxetine for ADHD^5^. Worryingly, many major pharmaceutical companies are decreasing their research efforts and investment in this area^6^. This apparent stagnation in progress is the result of a lack of understanding of the pathogenesis of these conditions, which hinders the identification of novel targets for drug discovery^7^, and also the limitations of current diagnostic categories in defining mechanistically discrete disorders^8^. A route to address these limitations involves integrating biological data at scale and across, rather than within, diagnostic classifications^9 10^. Research conducted in this manner can explore the aetiological and biological commonalities between diagnoses revealed by genetic studies^11^, accelerating discovery on complex disorders and informing novel therapeutic strategies, pharmacological and non-pharmacological, firmly grounded in biology^12^.

Recent large-scale studies have built on the hypothesis that psychiatric phenotypes do not always reflect distinct underlying pathogenic processes and that some genetic risk factors are shared between neuropsychiatric disorders^13-15^. This echoes the widely acknowledged clinical observation that many symptoms are features of multiple disorders and that patients often challenge current diagnostic classifications by presenting with characteristics of more than one disorder^16^. What is currently not known, however, is to what extent this distribution of cross-disorder symptoms is related to the shared genetic liability between neurodevelopmental conditions^15 17^. Commonalities in genetic risk factors might help identify a shared underlying biology, but this line of inquiry cannot be pursued without well-characterised cross-disorder samples, scarce even within large international consortia. In fact, it has been explicitly suggested that the majority of samples used in published genetic discovery studies have not been collected with the required amount of phenotypic data necessary to advance diagnostics, stratification and treatment^18^. Thus, many research groups have directed their efforts to access resources with large amounts of routinely collected data, such as population biobanks and electronic health record systems, from which rich phenotypic data can be derived^18-20^. However, some common limitations of these include selection biases and underrepresentation of clinically severe disorders^20 21^. These can be exemplified by a recent genetic study on 106,160 patients across four US healthcare systems, where only 522 individuals with a ICD-9/10 diagnosis of schizophrenia were included^22^. Such is a classic quandary in psychiatric genomics^23^, in which the setup of research studies leads to either a large case sample with minimal phenotyping or an extensively phenotyped one with fewer individuals.

The *Digital Repository for Amalgamating GenOmic and Neuropsychiatric Data* (DRAGON-Data) was therefore established at Cardiff University as a means of developing a platform where cross-disorder analyses of large well-phenotyped samples are possible. This approach integrates multiple existing case datasets with genetic, clinical, environmental, and developmental data. The focus on mental health across disorder boundaries and at scale aims to improve understanding of the pathophysiology of adult and child-onset neurodevelopmental and psychiatric disorders, providing opportunities to combine diagnosis-led and symptom-led research. DRAGON-Data shares a focus with previous initiatives to collate psychiatric phenotype data, which have included the Genetics of Endophenotypes of Neurofunction to Understand Schizophrenia (GENUS) consortium^24^, the International Consortium for Schizotypy Research (ICSR)^25^, the International 22q11.2 Deletion Syndrome Brain Behaviour Consortium (22q11.2DS IBBC)^26^, the Psychosis Endophenotypes International Consortium^27^, the Genes to Mental Health (G2MH) network, and ongoing efforts to collate phenotype data within the Psychiatric Genomics Consortium (PGC)^28^. However, all these projects have typically focused on a single disorder or group of closely related disorders, while DRAGON-Data seeks to integrate data from a range of psychiatric disorders across the symptomatology and developmental continua.

The current paper describes the formation of DRAGON-Data through the curation and harmonisation of phenotypic and genetic information across existing cohorts. This process has been informed by a series of legal and ethical considerations on the evolving landscape of individual-level data sharing, which is required to ensure the sustainability of this repository as a resource for current and future researchers. Therefore, the governance framework of DRAGON-Data is also described, which enables the access and reuse of its data in ways that align with confidentiality regulations and the ethics of participating studies.

## Methods and Analysis

### Studies included

Fifteen studies from the MRC Centre for Neuropsychiatric Genetics and Genomics at Cardiff University (MRC CNGG; https://www.cardiff.ac.uk/mrc-centre-neuropsychiatric-genetics-genomics) were included in this project. A summary of the studies can be found in **Table 1**. Each study had its own approved research ethics, whilst ethical approval for the curation and development of DRAGON-Data was obtained from Cardiff University’s School of Medicine Research Ethics Committee (Ref: 19/72). The studies included participants who were adults with psychiatric disorders, children (defined as up to age 16 or age 18) with neurodevelopmental disorders, children of parents with psychiatric disorders, and both children and adult carriers of rare neurodevelopmental risk copy number variants (ND-CNVs).

**Table 1.**
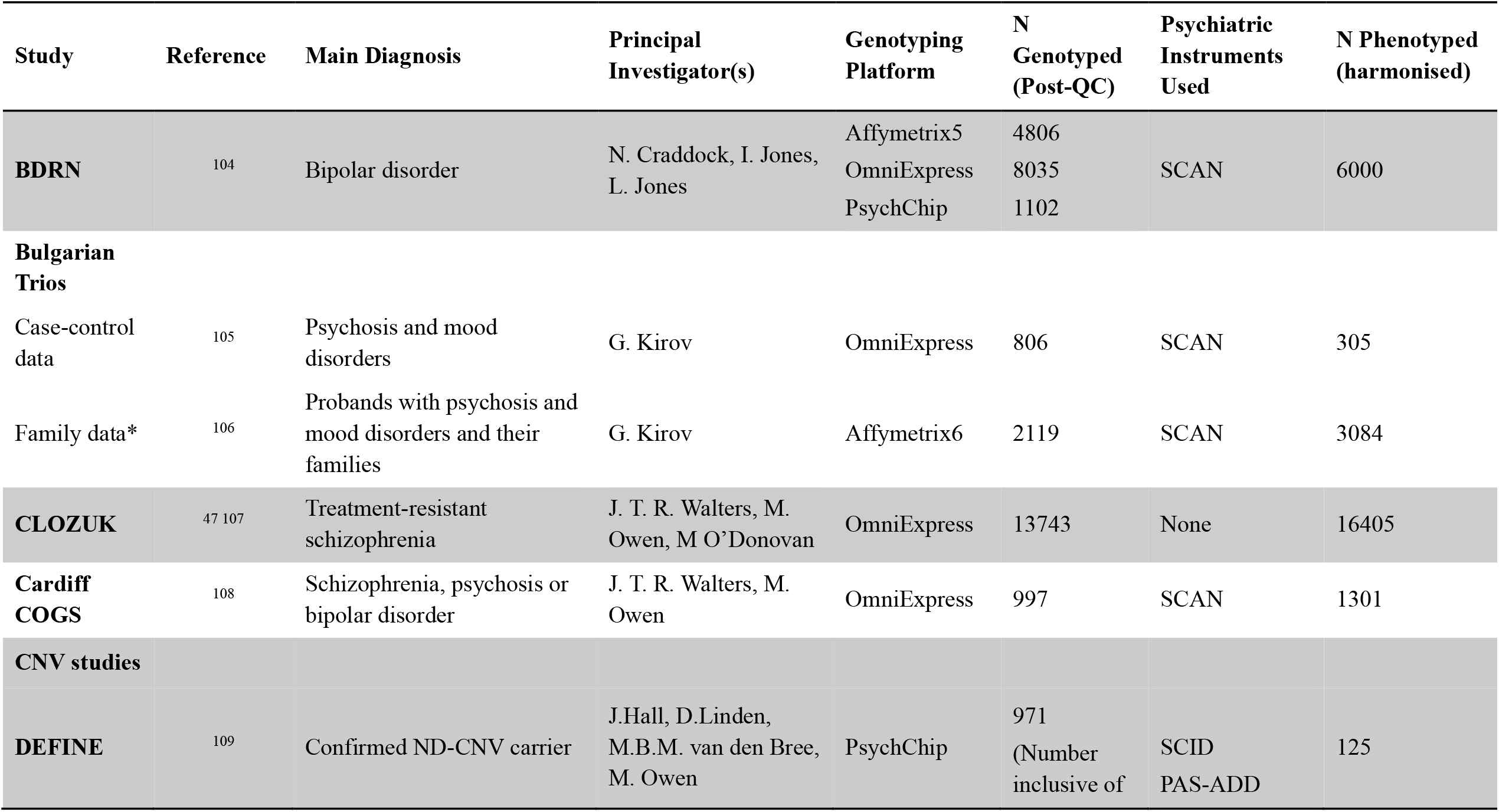
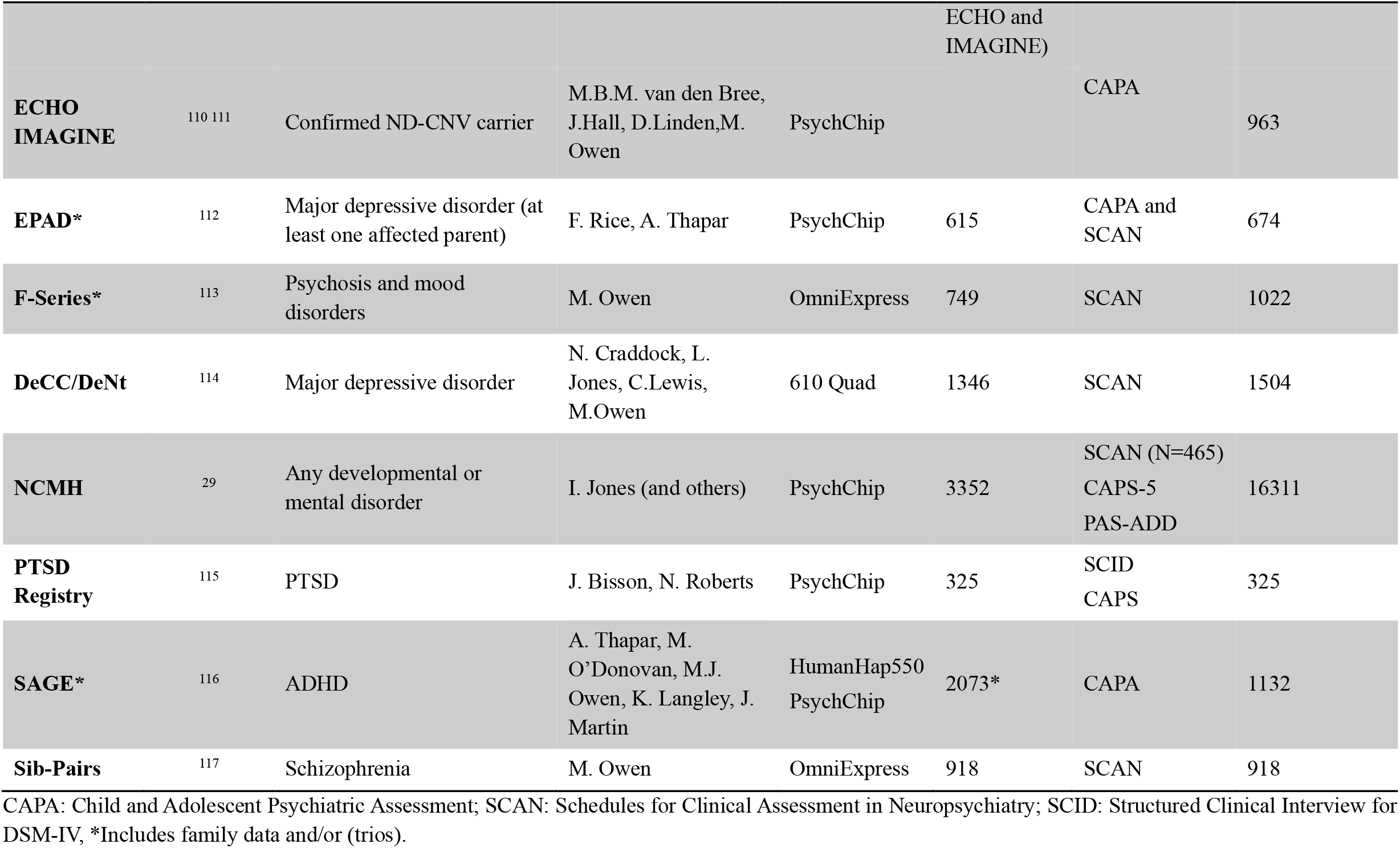
Studies included in DRAGON-Data.

### Phenotypic data harmonisation strategy

The process of curating the phenotypic data is outlined in Figure 1. Initially, investigators from all studies completed a proforma detailing the data and types of measures available, including the study clinical interviews, rating scales and self-report questionnaires. All but one of the studies included a structured clinical interview, and thus consistent symptom-level data were available (Error! Reference source not found.), along with a detailed phenotype data.

**Figure 1.**
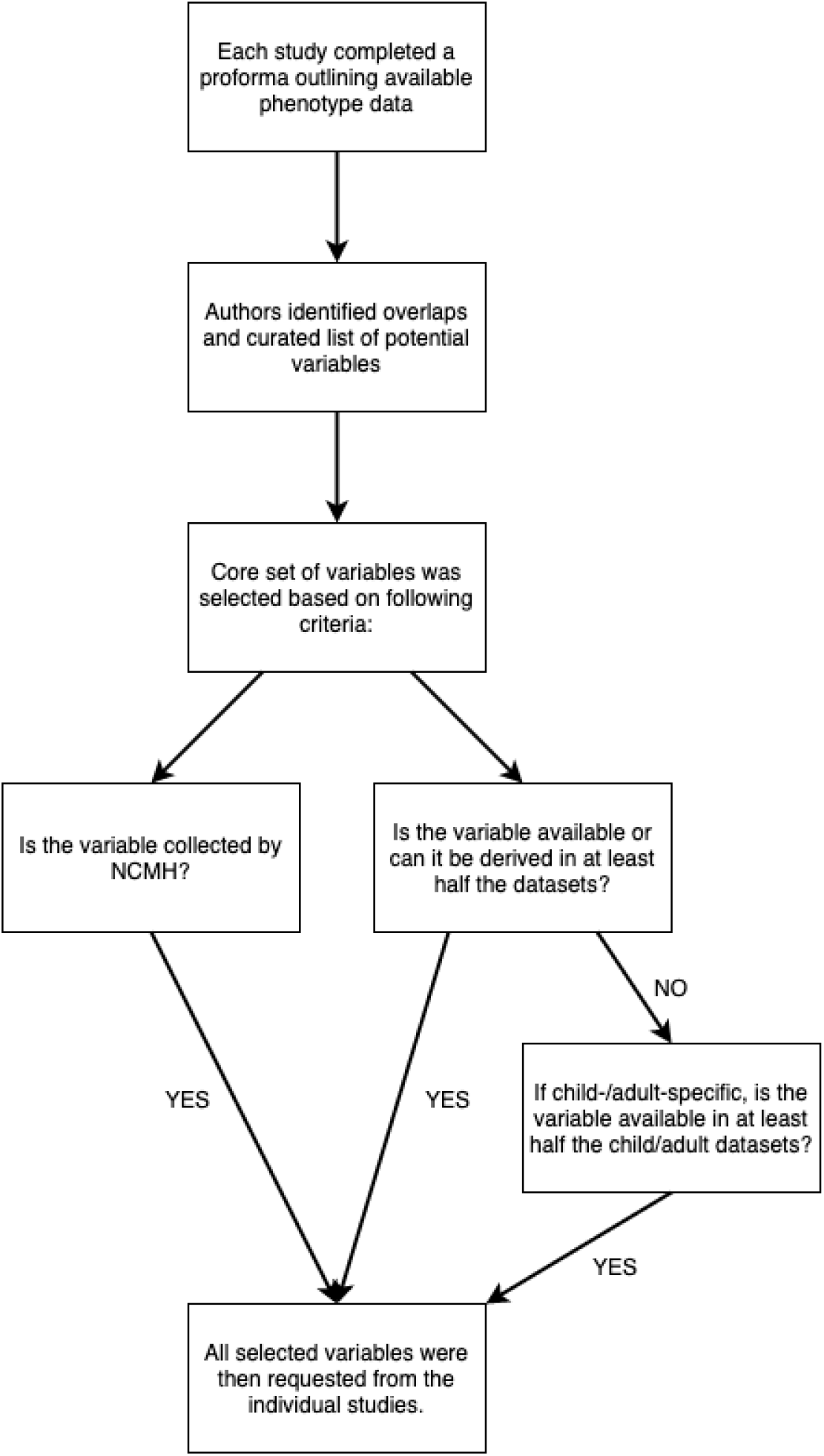
Curation of phenotypic data.

We compared all the variables to identify overlaps and resolve situations where the same information might have been differently labelled across studies. We also defined a core set of variables (**Table 2**), focused on information relevant and applicable to cross-disorder research. A primary consideration for including a variable among this core set was whether it was collected as part of the National Centre for Mental Health (NCMH) research programme. The NCMH is a Welsh Government-funded research centre that investigates neurodevelopmental, psychiatric and neurodegenerative disorders across the lifespan. Its cohort is the largest sample with phenotype data available to us, and a cross-disorder resource in itself^29^. As NCMH is still being expanded by recruitment of participants, maximising its compatibility with DRAGON-Data was desired. Additionally, every core variable was required to be available in at least half the current datasets, taking into consideration that some data might be specific to child or adult cohorts. Variables that were not available in NCMH and were present in less than half the studies were only included if they could be derived from existing data to achieve the representation threshold.

**Table 2.** List of phenotypic variables included in DRAGON-Data.

| **Variables Included** | **Number of studies** | **Number of participants** |
| --- | --- | --- |
| **Symptoms** |  |  |
| Depression | 12 | 15410 |
| Mania | 11 | 13906 |
| Psychosis | 9 | 12072 |
| ADHD | 4 | 2460 |
| Anxiety | 4 | 2478 |
| Conduct disorders | 4 | 2460 |
| Autism | 4 | 2460 |
| PTSD | 1 | 325 |
| **Treatment history** | 13 | 31164 |
| **Clinical / illness history** |  |  |
| Age of onset | 10 | 29023 |
| Hospital admissions | 7 | 26372 |
| Suicidal ideation | 12 | 15410 |
| **Adverse life events** | 6 | 9594 |
| **Education** | 9 | 24790 |
| **Substance use** | 13 | 29997 |
| **Family history of psychiatric illness** | 8 | 21473 |
| **Physical health** | 11 | 27725 |
| **Functioning** |  |  |
| Standardised measure of functioning (e.g. Global Assessment Scale) | 5 | 6260 |
| Marital / relationship status | 7 | 23290 |
| Current occupation | 7 | 25597 |
| **Cognitive function** | 7 | 5048 |

Number of participants refers to the number of data points available for each set of variables listed.

### Challenges of harmonising phenotypic data

#### Measuring and rating psychopathology

The individual studies that form DRAGON-Data were designed using standard protocols for psychiatric research, and collected similar phenotypic data. However, they also used a range of different interviews, rating scales and questionnaires.This creates well-known challenges for data harmonisation^30 31^. In general, it should be noted that caution has to be exercised when amalgamating data from different studies even when these claim to use the same measures. Potential differences can include:

- Versioning: Measures can differ considerably between versions, with items being added or removed and definitions changing.
- Rating definitions: Ordinal scales can be named (e.g. 1=“mild”, 2=“moderate”, etc) resulting in a categorical or integer variable depending on study protocol. Some scales (e.g. OPCRIT^32^) can include items for which decimal point rating is acceptable, which could be transformed into continuous variables.
- Rating timeframes: Symptom and event data can be evaluated over different timeframes spanning weeks, months or years; and recorded as current, worst or lifetime occurrences. When integrating adult and childhood studies, it should be considered that events defined for the “lifetime” are not directly comparable due to intrinsic differences in this period of assessment. Measures that evaluate personality and behavioural traits might also not be completely consistent given the changes in these throughout the lifetime^33^.
- Sources of information: A difference between adult and childhood studies is that the latter is more likely to use multiple informants (participants, their siblings, parents and teachers). Harmonising all these reports can be difficult and might also require a prior compatibility assessment^34^.

The considerations above apply to individual studies, but they can add particular difficulty to reflect complex outcomes in a larger harmonised dataset. As an example, we highlight the different ratings of suicidal ideation across the DRAGON-Data studies (**Table 3**). Note that these studies differed in whether they considered single versus multiple suicide attempts, duration of suicidal ideation or seriousness of attempts. This is likely to reflect the existence of different definitions of suicidal behaviour used in different research contexts^35 36^, and illustrates one of the challenges that can be faced when merging data from different studies.

**Table 3.** Rating scales for suicidal ideation across the studies.

| **Study** | **Suicidal Ideation: Rating Scale** |
| --- | --- |
| **CoMPaSS** | 0. Absent<br>1. Tedium Vitae<br>2. Suicidal Ideation<br>3. Attempt unlikely to result in death<br>4. Attempt likely to result in death<br>5. Multiple attempts likely to result in death |
| **NCMH** | 0. Absent<br>1. Tedium vitae<br>2. Suicidal ideation<br>3. Attempt unlikely to result in death<br>4. Attempt likely to result in death<br>5. Multiple attempts unlikely to result in death<br>6. Multiple attempts likely to result in death |
| **ECHO, IMAGINE, SAGE & EPAD (children only)** | Binary variables (yes/no) covering:<br>• Thoughts about death or suicide<br>• Suicide attempts<br>• Non-suicidal self-harm |
| **EPAD (parents only)** | Suicide attempt or self-harm:<br>1. Mild<br>2. Moderate<br>3. Severe |
| **PTSD Registry** | Question covers suicide attempts and self-harm in the context of borderline personality disorder:<br>1. Inadequate information<br>2. False or absent<br>3. Sub-threshold<br>4. Threshold or true |
| **Sib-Pairs & F-series** | 0. None<br>1. 1 week duration or one attempt<br>2. 2 weeks duration<br>3. At least one month |
| **Bulgarian Trios (family and case data)** | 0. Not present<br>1. Thoughts but no attempts<br>2. Attempt at suicide<br>3. Serious attempt |
|  | 4. Multiple serious attempt |
| **BDRN** | Suicidal ideation:<br>1. Yes<br>2. No<br>3. Unknown |
| **DeCC and DeNt** | 1. Deliberately considered but no attempt<br>2. Injured self or made attempt but no serious harm<br>3. Suicide attempt resulting in serious harm<br>4. Suicide attempt designed to result in death<br>5. Uncertain |

Note: No variable for suicidal ideation or attempts in DEFINE

#### Sampling from the Population

Recruitment strategies and inclusion criteria can affect the characteristics of the samples, creating differences between them and making them unrepresentative of the population from which they are drawn. It has been suggested that participants enrolled in research studies of serious mental illness display better functional outcomes than are typical for those with the disorders in the wider population^37^, when compared against naturalistic samples from outpatient services^38^. Population cohort studies also suggest that those with more severe psychopathology and higher genetic loading for psychiatric disorder are more likely to drop out, leading to underrepresentation particularly in longitudinal samples^39^. Media used to approach these participants also play a role in the sample characteristics, with internet-based recruitment engaging larger proportions of highly-educated female individuals but also those from ethnic minorities^40 41^. For most studies in DRAGON-Data, recruitment was based on clinically ascertained, prevalent cases and therefore are likely to have over-sampled participants with severe, chronic illness and under-sampled individuals who recovered and/or were discharged from services. Additionally, in common mental health conditions such as depression and anxiety, this might also over-represent women who are more likely to access help than affected males^42^. A special case in terms of sample composition also concerns the DEFINE, ECHO and IMAGINE studies, which specifically focused on carriers of ND-CNVs. Including these samples has important implications for research examining genotype-phenotype associations in the combined dataset, as improperly accounting for their genotype-led recruitment might bias calculations on the prevalence of genetic or environmental risk factors. However, they are important to integrate as they also enable comparative research into the role of these in people with and without highly penetrant genetic variants^43^.

#### Study Protocol

Samples were recruited following longitudinal and cross-sectional designs. The existence of a follow-up period in longitudinal studies establishes a temporal order for symptom and event measures, which provides another level of detail over the broader definitions found in cross-sectional designs. The cross-sectional studies collected a mixture of current, worst episode and lifetime symptom measures. As it has been previously described in the context of causal inference^44^, it is not advisable to combine longitudinal measures into or with “lifetime ever” variables, since this assumes that the events they reflect did not occur outside of the study assessment periods. Other issues that can affect the compatibility of different designs are attrition (in longitudinal studies), participant issues in completing assessments (e.g. length of time required) and the mode in which the study was conducted. Within DRAGON-Data most studies were conducted face to face with participants before the onset of the COVID-19 pandemic, but also utilised telephone interviews, postal questionnaires and online data collection. This could affect how questions are interpreted and in turn, the likelihood and content of participants’ responses. In addition, there is evidence that participants may be more willing to disclose sensitive information in some settings than others^45 46^.

#### Diagnosis

Due to the different focus of individual DRAGON-Data studies, there were differences in the ways that diagnoses were made. Most studies used standardised interviews and medical records (where available) to derive consensus research diagnoses, with CLOZUK validating their ascertainment (based on intake of the antipsychotic clozapine) against research interviews^47^. The NCMH population sample used self-report, asking participants to report diagnoses that they had been given by a health professional. This is an approach taken by other large studies such as the UK Biobank^48^. While data obtained via self-reports can be of poorer resolution than that from a structured interview, this approach has the advantage of allowing faster recruitment of larger samples^49^. The accuracy of self-report diagnoses needs also to be considered, which may differ by diagnosis. Self-reported diagnoses of specific, chronic mental health conditions that require involvement with secondary psychiatric services, such as schizophrenia, may be more accurate than reports of common mental health conditions, such as depression, that typically encompass a wide range of presentations and can be diagnosed and treated in a variety of health settings. This can introduce variability in defining phenotypes with impacts on study results. Research attempting to estimate the heritability of depressive disorders using inconsistent diagnostic criteria classically demonstrated this^50^; and recent work employing samples with broad, self-report definitions of depression to identify genetic risk loci have also resulted in signals that are not specific to this condition^51^. To ameliorate these problems, the studies included in DRAGON-Data have focused on categorical diagnoses rated according to the Diagnostic and Statistical Manual of Mental Disorders (DSM) or International Statistical Classification of Diseases (ICD) criteria. While these are standard criteria, it has been proposed that a better approach to diagnostic classification may be to focus on dimensional measures of psychopathology, such as the National Institute for Mental Health’s Research Domain Criteria (RDoC^52^). This approach may be adopted in the future as it could facilitate combining datasets to conduct cross-disorder research, given that many symptoms overlap diagnostic boundaries, such as the overlapping mood and psychotic symptoms observed in both schizophrenia and bipolar disorder^8^.

#### Key Recommendations

Based on our experience developing DRAGON-Data, we suggest some recommendations for the harmonisation and analysis of clinical data:

- Consider the broad research questions that can be addressed with the creation of a clinical database. Consult with principal investigators and field researchers to identify the variables that will be needed to address these aims.
- Identify measures (e.g., questionnaires and interviews) that are in common across the datasets included. These measures may be easier to harmonise for analysis, though the factors outlined above should be considered to ensure comparability.
- Record accurate information about each study variable including measure used, version number, rating definitions, rating timeframe and source of information. This aids in the identification of comparable variables.
- Where new (secondary) variables have been derived from others, and are designed to be comparable, information should be recorded about the (primary) variables used from each study to derive those secondary variables.
- A comprehensive data dictionary should accompany the database that incorporates the information outlined above. At a minimum, each variable should have recorded: name, description, definition and coding of missing values. Within the data dictionary, variables should be highlighted if they are in common across the datasets, as these may be suitable to analyse together. It is noteworthy that this curation and creation of dictionaries may often need to occur after the data collection, so researchers and funders should allow sufficient staff resources for the accurate completion of this task.
- Include basic demographic information to evaluate the representativeness of the sample, including age range, sex, ethnicity and education.
- Datasets do not need to be combined into a single data file. A database that houses the datasets and allows an easy combination of selected studies and variables avoids the need for a single, large-scale dataset and minimises the computational requirements for the querying and extraction of data.

### Genetic data harmonisation strategy

#### Format and genome assembly standardisation

We developed an in-house genotype quality control (QC) pipeline to facilitate standardised procedures for all aspects of genetic analysis (**Figure 2**), available at https://github.com/CardiffMRCPathfinder/GenotypeQCtoHRC. The pipeline begins with conversion of genotype data into binary PLINK format^53 54^. Genotyping platform was inferred by comparing chromosome and basepair positions of the genotypes on each dataset and 166 array manifests^55^. Across the datasets in DRAGON-Data, Illumina chips are by far the most common (**Table 1**).

**Figure 2.**
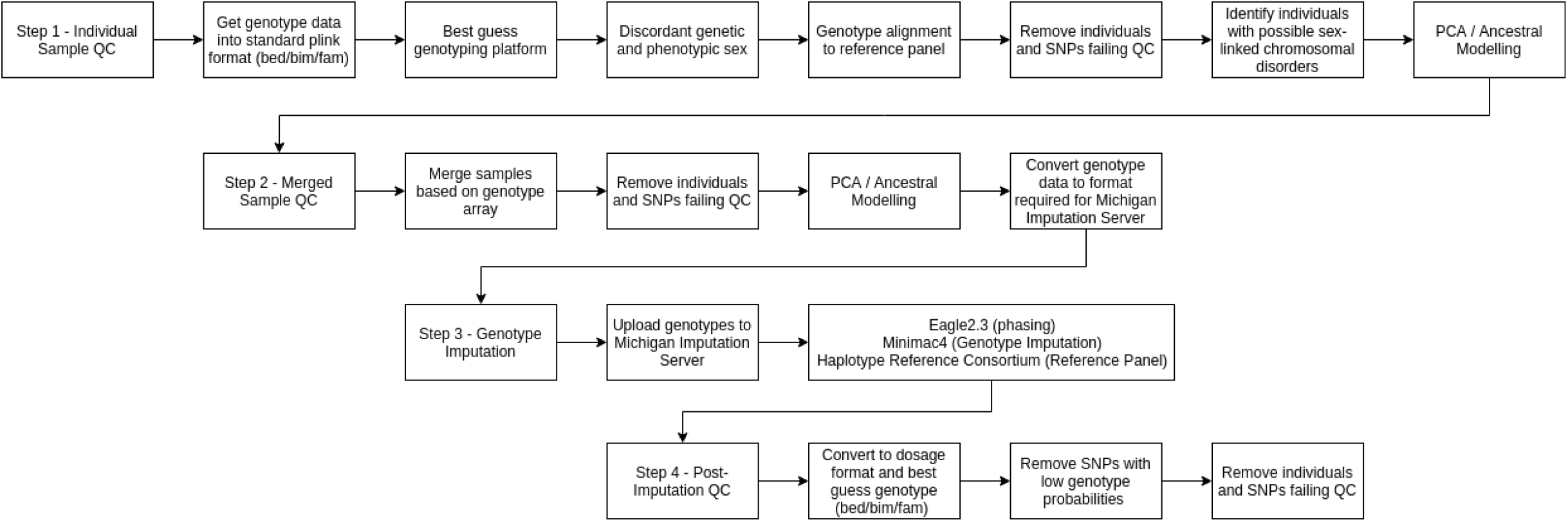
DRAGON-Data pipeline for SNP genotype QC and imputation.

To maximise the number of SNPs available for imputation, we performed alignment of local genotype data against the Haplotype Reference Consortium (HRC) panel v.1.1^56^ using Genotype Harmoniser v1.42^57^. Genotype Harmoniser is a Java-based application that compares SNP information in the user data against a reference dataset such as an imputation panel. Where discordant SNP information is present, for example due to allele mismatches, strand flips or different SNP identifiers, the user genotype data is updated to match that of the reference panel. We have observed that differences in genome build between the original and reference dataset result in Genotype Harmoniser discarding large numbers (e.g. more than 50%) of the original SNPs. If present, instances of this behaviour are flagged by our pipeline and solved via a local implementation of the widely-used Liftover Tool^58^ to retrieve physical coordinates in the appropriate b37/hg19 format.

#### Sex-based quality control

We performed checks for discordant phenotypic and biological sex using the “sex-check” function in PLINK v1.9. This function is reliant on the presence of at least one sex chromosome. Discordant findings in the absence of complementary information from the individual (e.g. a disclosure of gender transitioning) are suggestive of either a sample mix-up during genotyping or an inaccurately recorded phenotype. If no resolution can be reached these samples are excluded from further analysis. Where no sex information is present in the original dataset, the sample is retained. If genotype calls from both sex chromosomes are present, call rates at the Y chromosome are used to assess the presence of individuals with sex-linked chromosomal disorders such as Turner (X0) or Klinefelter (XXY) syndromes^59^. Individuals with suggestive sex-linked chromosomal disorders are flagged for further investigation.

#### Call-rate quality control

We removed SNPs with low call rates (<0.95), individuals with low genotyping rates (<0.95), markers that fail the Hardy-Weinberg Equilibrium test (mid-p<10^−6^) and those with a minor allele frequency (MAF) < 0.01. Duplicated individuals were removed unless they belong to known monozygotic twin pairs; however, first degree relatives are retained for studies with trio or family designs. This is the final step of the pre-imputation QC. Afterwards genotypes are converted to VCF format using PLINK, sorted using vcftools v0.116^60^ and compressed to .gz format.

#### Assessment of population structure

While not strictly part of a QC process, the generation of principal components (PCs) using genotype data is needed to identify and account for population and ancestral substructures that can bias the results of association studies^61^. Our pipeline addresses this by generating PCs using the GENESIS suite, implemented in R. Within it, the PC-AiR^62^ function allows us to process both unrelated and family-based datasets, as it accounts for known or cryptic relatedness via the calculation of genotype relatedness matrices (GRMs). PCs generated by this method can readily be used to correct for population structure in regression-based analyses.

A more detailed ancestry analysis is also performed on each dataset, following a similar procedure to that described in Legge et al. 2019^63^. First the available SNPs are restricted to those on the set of 167 ancestry informative markers (AIMs) contained in the EUROFORGEN^64^ and 55-AISNP^65^ forensic panels, many of which are common across the different Illumina genotyping platforms. Afterwards, the dataset is merged with a public reference panel with known ancestries, a combination of the Human Genome Diversity Project (HGDP)^66^ and South Asian Genome Project (SAGP)^67^ datasets. This reference contains 1108 samples from 62 worldwide populations, which have been subdivided in 7 biogeographical ancestries^68^ (“Subsaharan African”, “North African”, “European”, “Southwest Asian”, “East Asian”, “Native American” and “Oceanian”). In order to perform the ancestry inference, a number of PCs, determined using the Tracy-Widom test for eigenvalues^61^, are then derived solely on the reference panel, and a prediction model is trained using Fisher’s Linear Discriminant Analysis algorithm. The samples with unknown ancestries are then “projected” onto the reference panel PCs^69^, and their ancestry is estimated using the prediction model. At least 80% probability of a given ancestry is required to automatically assign an individual to it, though the admixture patterns of individuals not achieving this probability can still be manually examined.

#### Genotype imputation

The Michigan Imputation Server (MIS) is a cloud-based resource that facilitates haplotype pre-phasing and genotype imputation^70^. The MIS also houses the HRC panel, containing genotypes of over 60,000 individuals across multiple ancestral backgrounds^56^. There are substantial improvements in imputation quality using the HRC reference over 1000 genomes, particularly at lower MAF thresholds^71^. The MIS also performs some SNP quality control before phasing, including removal of SNPs if they contain irregular allele codes, duplicate IDs, indels, monomorphic SNPs, discordant alleles between the user and population reference panel alleles and low call rates of < 0.9. Though other options are available, our dataset is processed via Eagle v2.3 pre-phasing^72^ and MiniMac3 imputation^70^ using HRC v1.1 as the reference panel.

After genotype imputation, imputed data is stored in .vcf.gz format, with accompanying info files containing information about the quality of imputed variants. Data is converted into .pgen format using PLINK v2 and subsequently into standard .bed/.bim/.fam format. Specifically, we remove SNPs where individual genotype probabilities are < 0.9, MAF <1%, genotyping rate < 0.95 and hwe < 1E-4. SNPs can be extracted at various imputation quality thresholds (R2). A conversion to best-guess genotypes is also performed in PLINK v2, after applying imputation quality thresholds (INFO < 0.3).

#### Copy Number Variant Calling

Most of the samples in DRAGON-Data include raw genotype information, enabling us to perform copy number variant (CNV) calling. We developed an in-house CNV QC pipeline to facilitate standardised procedures for all aspects of this procedure (**Figure 3**), available at https://github.com/CardiffMRCPathfinder/NeurodevelopmentalCNVCalling.

**Figure 3.**
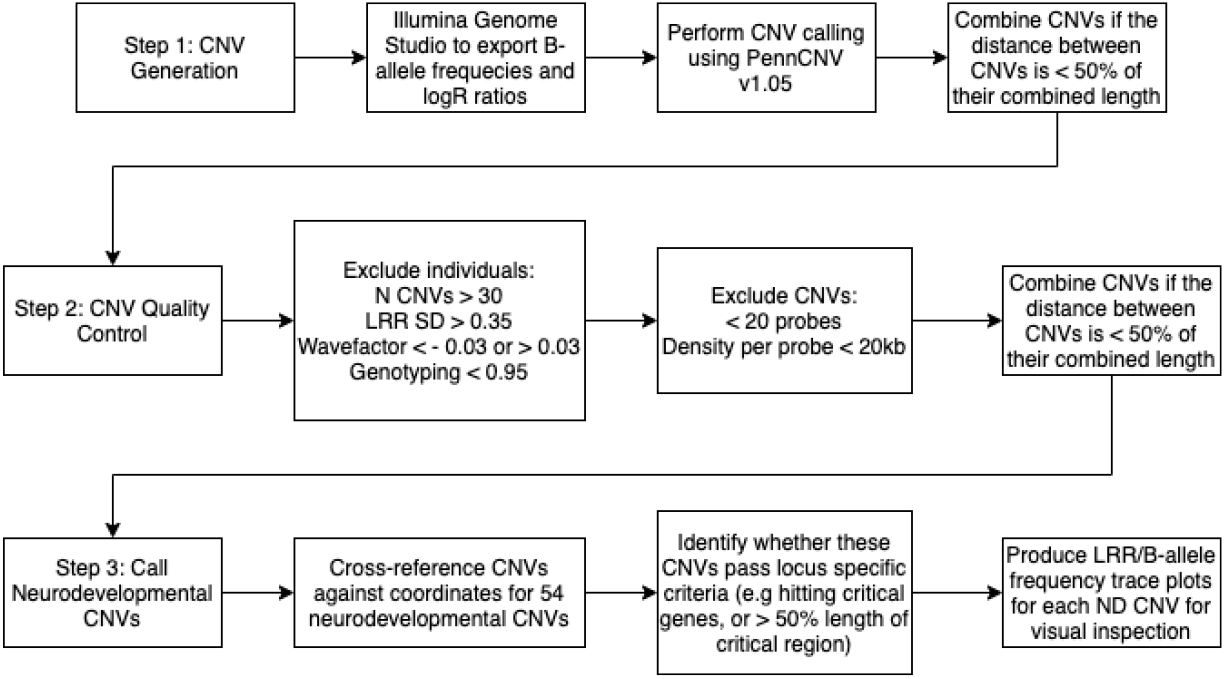
DRAGON-Data pipeline for CNV Calling.

First, we extract b-allele frequencies and logR ratios for each sample using Illumina GenomeStudio v2.05. CNV calling is performed using PennCNV v1.05 with genomic control correction^73^. CNVs are subsequently merged if the total distance between CNVs is less than 50% of their combined length. Appropriate PFB and GC content files are generated as recommended by PennCNV. Filters are applied to remove CNVs with QC fewer than 20 probes, less than 20KB in length or with confidence scores < 5. Individuals are excluded if they have more than 30 CNVs, large logR ratios > 0.35 or high or low wavefactor (less than -0.03 or greater than 0.03), however these parameters should be modified depending on the genotyping platform used.

Initially, CNVs called using this pipeline are cross-referenced against a list of 54 pathogenic CNVs known to confer increased risk of schizophrenia, autism, intellectual disability and major depressive disorder^74^. There are several advantages to prioritising these CNVs: First, they are typically large (>100KB) and are more reliably called across different genotyping platforms. Second, these CNVs are pleiotropic and lack complete penetrance for specific disorders meaning they are good candidates for investigating associations with psychiatric cross-disorder phenotypes.

### Challenges of harmonising genetic data

#### Genotyping arrays and genomic assemblies

In DRAGON-Data, a variety of genotyping arrays were used both within and between studies. This presents challenges for merging and imputing datasets. All the genotyping arrays analysed have a large set of common variants (a “GWAS backbone”), with most differences due to the inclusion of custom markers tagging rare exonic variation. The accuracy of genotype imputation is improved with larger sample sizes, plateauing around 2,000 samples^75^, though there must also be sufficient numbers of genotyped markers (at least 200,000 SNPs^76^) that overlap with the imputation reference panel after genotype quality control. We, therefore, grouped datasets that were genotyped on the same, or similar arrays. This resulted in four separate imputation batches for samples genotyped on the OmniExpress, PsychChip/Illumina HumanCoreExome, Illumina 610 Quad/Illumina HumanHap550 and Affymetrix5 platforms.

We observed substantial batch effects in the pairwise comparison of samples after undergoing routine QC. Further inspection of the data revealed this was caused by palindromic SNPs (AT/TA or CG/GC genotypes), which resulted in erroneous allele frequencies which differed across datasets when the minor allele frequency was high (> 0.4). This issue was only apparent after merging datasets, which mirrors the experience of the eMERGE consortium^77^. Removal of these SNPs resulted in the loss of obvious batch effects across the first 10 PCs tested.

#### Identifying duplicate samples

It is not uncommon for the same individual to be recruited into more than one psychiatric research study. Unless the individual voluntarily reports they have participated in a known existing study, this information would not be known to researchers in other groups. We identified 1909/41957 duplicate individuals (4.5%) across the entire dataset using genetic relatedness checks and retained the sample with the highest number of high quality imputed markers. In total,

#### Processing of public GWAS summary statistics

When performing genetic analyses such as polygenic risk scoring, LD score regression or other analyses, multiple GWAS summary statistics are required. Despite some proposals for standardisation^78-80^, the output from GWAS software is still highly variable and lacks even consistent headings across individual studies. Processing of these files is thus not user-friendly, typically requiring manual curation, for example filtering by imputation quality, allele frequency or changing header names to match the required format of specific programs. To address these issues, we developed an R pipeline (summaRygwasqc) that automatically processes GWAS summary statistics files and performs quality control filtering, aligns SNP information against the HRC reference panel and converts summary data to a standardised format that is compatible with PRSICE2^81^, PRScs^82^ and LDSC^83^ (**Figure 4**). This code is available at https://github.com/CardiffMRCPathfinder/summaRygwasqc.

**Figure 4.**
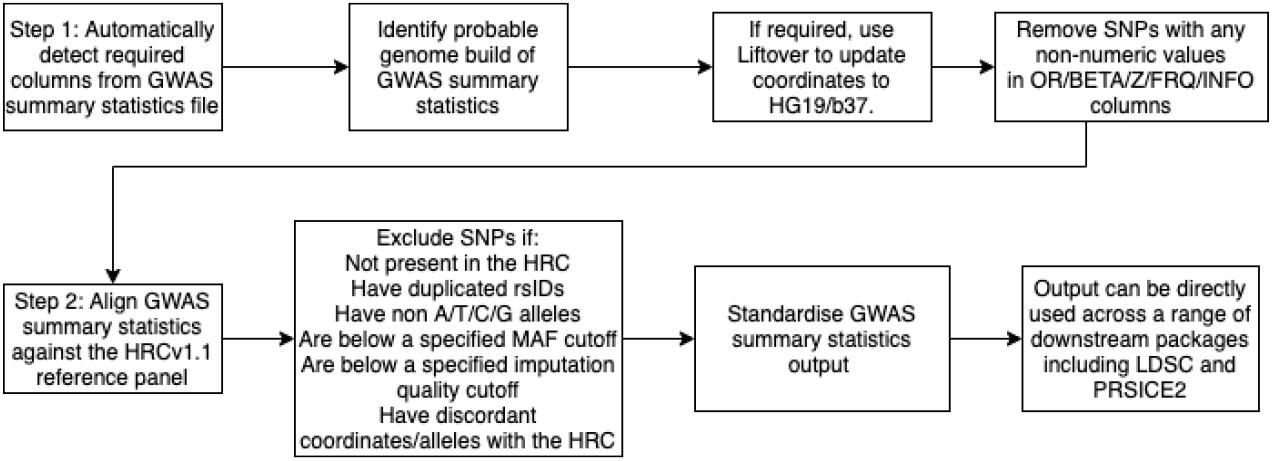
DRAGON-Data Pipeline for standardising genome-wide association summary statistics (summaRygwasqc)

#### Key Recommendations

Based on our experience developing DRAGON-Data, we offer some recommendations for the amalgamation and analysis of genomic data across multiple studies:

1. Imputation should only be performed on samples that have been genotyped on the same array type, or where there is substantial SNP overlap after QC. Furthermore, when performing QC after imputation, removal of palindromic SNPs with high MAF (>0.4) is essential to minimising batch effects for samples genotyped on different arrays.
2. When analysing CNV data across arrays, due to potential differences in probe density and coverage, it is vital that plots such as those for b-allele frequency drift, number of CNVs called per individual and LogR ratio standard deviation are visually inspected to ensure the quality of the resulting calls.
3. Publicly available genome-wide association summary statistics should be examined, manually or through scripting, to ensure that their information can be processed in a coherent and standardised way in downstream analyses. At a minimum, most genomic analysis software requires a form of SNP name or identifier, chromosome number (CHR), basepair position (BP), allele code (A1/A2), association p-value and a metric for the association effect size (OR/logOR/beta/Z, which should always correspond with A1). Additional columns such as the allele frequency of A1, INFO/R2 imputation quality metrics and sample size columns can also be helpful.

### The DRAGON-Data harmonised dataset

**Table 2** displays an overview of the variables held by each study included in the final DRAGON-Data data freeze. A full list of the variables included in DRAGON-Data can be found in **Supplementary Table 1** although the exact variables included varied between studies. All the studies except CLOZUK included a semi-structured clinical diagnostic interview, most commonly the Schedule for Clinical Assessment in Neuropsychiatry (SCAN^84^) for adults and the Child and Adolescent Psychiatric Assessment (CAPA^85^) for children and adolescents. Twelve of the fifteen studies collected data on individual symptoms. The NCMH study includes a brief assessment that does not include questions about individual symptoms, although a subgroup of this sample (n=485) has completed more detailed interviews that include symptoms. The most common types of symptoms covered across all studies were depressive, manic and psychotic symptoms. Aside from symptoms, other variables with good coverage across studies were lifetime history of treatment (13/15), substance use (13/15) and history of suicidal ideation and attempts (12/15). The demographic characteristics of the studies are shown in **Supplementary Table 1**. The harmonised phenotype data is stored in a pseudonymised format within a secure database. There is an accompanying data dictionary cataloguing all available variables with names, descriptions and ratings and cross-referencing of comparable measures across the studies.

### Joint genetic-phenotypic data analysis

All the DRAGON-Data data have been securely stored in HAWK, a high-performance computing (HPC) cluster supported by the Supercomputing Wales infrastructure^86^, which comprises a network of 13,000 computer nodes distributed across four universities (Cardiff, Swansea, Bangor and Aberystwyth). This system allows the backed-up storage of genetic and phenotypic files, and their secure access by authorised users. Analysts in charge of curating genetic or phenotypic data are by default part of a “core project team” with unrestricted access to the entire DRAGON-Data, while data-contributing researchers are granted access to their own raw and curated data for any purpose. Undertaking cross-disorder analyses is facilitated through a framework by which any curator or data-contributing researcher can send a structured analytic proposal to the board of investigators, who then decides whether to grant access to the relevant data on purely scientific grounds. This is modelled after successful international consortia such as the PGC^28^, which in recent years has implemented responsible data sharing practices among hundreds of investigators.

There are two main approaches to analysing the data within DRAGON-Data: combining individual-level information from across the studies (“mega-analysis”) or through meta-analysis. While the latter is relatively straightforward, jointly analysing all samples allows for a better assessment of heterogeneity in the data and can increase statistical power^87 88^. However, combining samples is particularly problematic for the phenotypic data, as it requires recoding or modifying the variables to be comparable across studies, which could include deriving latent variables through factor analysis. Data combined in this way can be difficult to interpret due to the differences between studies outlined in the previous sections, and it is important to address this variability in both analytic techniques and interpretation of the results. Important considerations are whether the individual study variables are measuring the same construct and whether any variables derived from these are measuring the same construct as the original data. Note that none of these limitations applies to the genetic data, as (carefully) combining samples with large numbers of overlapping SNPs is a common procedure that is known to maximise both the number of successfully imputed variants and their quality^75 89 90^. Thus, the suitability of a mega-analysis or meta-analysis approach for studies using DRAGON-Data should be decided based on the availability, characteristics and biases of the phenotypic data.

Outside of the data quality control pipelines, genetic analyses in DRAGON-Data can be undertaken using other consolidated tools, such as PLINK^53^ or GCTA^91^. Responding to the rapid development of statistical methods to analyse complex phenotypes and “big data”, an effort has been made to integrate DRAGON-Data with the highly customisable R framework, via the use of data importers such as *GWASTools*^92^ and *bigsnpr*^93^. This allows using the approximately 1,700 tools currently offered by the Bioconductor suite^94^ in a large-scale genome-wide setting, and facilitates applying complex analytic techniques such as mixed-model regression^95^ and survival analysis^96^. Large-scale genomic storage solutions have not currently been implemented in DRAGON-Data, as the weak compression implemented in PLINK files and related formats allows for efficient querying of genotype data even in its imputed form^53,97^. However, these are active topics of research, and the upcoming development of the MPEG-G ISO standard will likely allow future data harmonisation initiatives to seamlessly incorporate whole-genome sequences^98^.

## Ethics and dissemination

### Governance

For studies to be incorporated into DRAGON-Data, the lead principal investigator needed to confirm approval from their institutional ethics committee. The protection and confidentiality of participant data were of the utmost importance throughout the design of DRAGON-Data and a number of safeguards were put in place to ensure the security, integrity, accuracy and privacy of participant data. Firstly, in line with the required safeguards for processing special category data stipulated in the EU General Data Protection Regulation (GDPR; Article 89)^99^, the principle of data minimisation was respected, with only limited individual-level data being requested from research groups. Furthermore, as a means of ensuring the confidentiality and privacy of participants, all data were pseudonymised, and no personal or phenotypic information that allowed individuals to be re-identified was retained. As genome-wide genetic information cannot effectively be anonymised without compromising its integrity^100^, all researchers accessing it must explicitly state that they will not attempt participant re-identification.

This project was conducted in line with Cardiff University’s Research Integrity and Governance Code of Practice, and ethical approval for the curation and development of the DRAGON-Data was obtained from Cardiff University’s School of Medicine Research Ethics Committee (Ref: 19/72). As described above, procedural safeguards were put in place to ensure secure managed access to the dataset through the HAWK system, with the most privileges restricted to the “core analyst team”. In addition, a process of oversight has been implemented for the approval of secondary research proposals, which are reviewed by the lead principal investigator of each contributing sample and must be approved before access to relevant, requested data can be granted. All genetic analyses carried out by secondary investigators also have to be carried out within the HAWK environment, which allows their monitoring and auditing to rapidly detect data misuses.

### Challenges of data sharing partnerships

The organisational challenges faced by DRAGON-Data highlight that potential data sharing requirements should be considered, as much as reasonably possible, at the outset of any research study. Studies will benefit from having a data sharing policy in place prior to the collection of any data as a means of maximising the value of collected data, increasing transparency and ensuring responsible future sharing of data. This will depend on sharing with whom, and for what purpose. Consent processes have changed dramatically over the last 30 years and historical studies will not all have explicit consent on the data sharing practices that are more commonly included today^101^. In certain situations, additional ethical permission may be required for data sharing when the sample is historical and or individuals can no longer be contactable. Thus, data sharing without that explicit permission can only occur within certain circumscribed situations.

When obtaining consent for future research, researchers should aim to be as inclusive as possible and allow participants to provide their written informed consent for general areas of research activity. In the context of broad consent, we would also advise the implementation of an oversight mechanism for the approval of future research studies. Participants entrust researchers to make reasonable decisions regarding future research on their behalf and the process of oversight adds further protection to participants, since not all future research uses can be predicted.

### Dissemination

At present, DRAGON-Data has been designed as a way of maximising the present and future utility of data collected at the MRC CNGG during the last thirty years. Given the complexity of the data, particularly the phenotypic portion, the first cross-disorder analyses of DRAGON-Data have been carried out by members of the core analytic team and the participating investigator groups. Results of these analyses will be shared through Cardiff University online data repositories and communicated through standard scientific channels such as peer-reviewed publications. Ultimately, through adapting the PGC open science model^102^ and taking advantage of the data-sharing frameworks supported by HDR UK, such as the DATAMIND Hub^103^, the DRAGON-Data resource will be available for external investigators where individual study consent and ethics permit such data sharing. This will ensure compliance with the permissions and ethics of individual studies, and will be based on the secondary analysis principles detailed in the Governance section.

## Data Availability

All code relating to the bioinformatics pipelines can be found in the data availability links.

https://github.com/CardiffMRCPathfinder/summaRygwasqc

https://github.com/CardiffMRCPathfinder/NeurodevelopmentalCNVCalling

https://github.com/CardiffMRCPathfinder/GenotypeQCtoHRC

