## Supplementary Table 1 for "DRAGON-Data: A platform and protocol for integrating genomic and phenotypic data across large psychiatric cohorts"

| Category | |
|---|---|
| **Variable Name** | **Study Information** |
| **Description of variable** | Age range, diagnosis |
| **CardiffCOGS** | 16+, schizophrenia, psychosis or bipolar disorder |
| **NCMH (brief assessment)** | 4+, history of developmental/mental disorder or intellectual disability, relatives included |
| **NCMH (indepth assessment)** | 18+, psychosis or mood disorder |
| **EPAD (child)** | 9-17, children of parents with depression |
| **EPAD (parent)** | 18+, history of depression, at least one child |
| **ECHO / IMAGINE (combined dataset)** | 2-18, confirmed CNV carrier |
| **DEFINE** | 18+, confirmed CNV carrier |
| **SAGE** | 6-18 years, diagnosed with or undergoing assessment for ADHD |
| **BDRN** | 18+, bipolar disorder |
| **Sib-Pair** | Individuals with psychosis and mood disorders |
| **F-Series** | Individuals with psychosis and mood disorders |
| **PTSD Registry** | 17+, individuals with PTSD |
| **Bulgarian Trios** | 13+, individuals with psychosis and mood disorders and their families |
| **Bulgarian Cases** | 21+, psychosis and mood disorders |
| **MDD Sample (Kings)** | 18+, major depressive disorder - recurrent |

| | | Identifiers | | | | |
|---|---|---|---|---|---|---|
| **Data Collection Methods** | **Symptom Data: Lifetime, Worst Episode or Current?** | **Cohort_ID** | **Participant_ID** | **DOB** | **Age_at_interview** | **Sex** |
| | | Identification code for the study the data is derived from | ID number for the individual participant | Month and year of birth | Age at interview | |
| SCAN + available clinical r | Lifetime | Sz | Study_ID | | m_ageinterview | m_sex |
| NCMH brief assessment (t | No symptom data | NCMH | Study_ID | | Age | Sex |
| SCAN + available clinical r | Lifetime | NCMH | Study_ID | | Age | Sex |
| CAPA | Ever and last three months | EPAD | ID | | age / Sage / Tage (offsp | Csex |
| SCAN | Current | EPAD | ID | | Mage, SMage, TMage, Fa | Gender |
| CAPA | Ever and last three months | ECHO | IDs | DOB | | |
| SCID | No symptom data | DEFINE | Participant_ID | | Age_at_Psychiatric_Inter | Sex |
| CAPA | Ever and last three months | SAGE | Study_ID | DOB | child_age | gender |
| SCAN + available clinical r | Lifetime | BDRN | StudyID | Month_and_year | Age_at_Interview | Sex |
| SCAN | Lifetime | Sz | Study_ID | | m_ageinterview | m_sex |
| SCAN | Lifetime | Sz | Study_ID | | m_ageinterview | m_sex |
| CAPS, PCL, SCID | Current | PTSD | Study_ID_numbe | DoB | Age | Gender |
| SCAN | Lifetime | Bulgarian Trios | SampleID | DOB | age | sex |
| SCAN | Lifetime | Bulgarian Cases | sample_id | DOB_proband | | sex |
| SCAN | Worst two episodes | Kings | MDD_subject_gl | Date_of_birth | Age_at_interview | Sex |

| | | Diagnosis | | | | | |
|---|---|---|---|---|---|---|---|
| **Ethnicity** | **Date_of_interview** | **ICD_1** | **ICD_2** | **ICD_3** | **DSM_1** | **DSM_2** | **DSM_3** |
| | | ICD-10 Diagnosis | ICD-10 Diagnosis | ICD-10 Diagnosis | DSM-IV Diagnosis | DSM-IV Diagnosis | DSM-IV Diagnosis |
| m_ethnicity | | ICD_Main | ICD_2 | ICD_3 | DSM_Main | DSM_2 | DSM_3 |
| Ethnicity | | Provisional_Diagnosis_1 / Provisional_Diagnosis_2 / Provisional_Diagnosis_3 / Provisional_Diagnosis_4 / P | | | | | |
| Ethnicity | | ICD10_Main | ICD10_2 | ICD10_3 | DSMIV_Main / D | DSMIV_2 / DSM | DSMIV_3 / DSM |
| Tmfamback / Tffamback - Mother and father's background | | | | | CADHD / cASD / CODD / CCD / Canx / cDD / cMD | | |
| Tmfamback / Tffamback - Mother and father's background | | | | | mdiagnosis / fdiagnosis (parent's diagnosis of depr | | |
| P_Fam_Back_1 | CAPAinterviewdate | | | | OCD / PanicDisorder / PanicWithAgoro / Agorophc | | |
| Ethnicity_ | Date_of_particpation_ | | | | Total_psychiatric_diagnoses / Any_Psychiatric_Dia | | |
| ethnicity_m / ethnicity_mg / ethnicity_f / | | See SAGE page for comprehensive list | | | | | |
| Ethnic_Origin | | ICD_Main | | | DSM_IV_Main | | |
| m_ethnicity | | m_ICDdiagnosis | | | m_DSMdiagnosis | | |
| m_ethnicity | | m_ICDdiagnosis | | | m_DSMdiagnosis | | |
| Ethnicity | Interview_Date | | | | DSM5PTSDdiagnosis | | |
| ethnic | | | | | DSM4 | | |
| | Date_collected | | | | DSM4 | | |
| Self_ethnicity | Consent_date | | | | | | |

| History of Symptoms | | | | | | | | |
|---|---|---|---|---|---|---|---|---|
| **Depression** | **Anxiety** | **Panic_Attacks** | **Mania** | **Psychosis** | **Dep_Dysphor** | **Dep_Irritability** | **Dep_Concentrati** | **Dep_Slowed_Activity** |
| Depression ever | Anxiety ever | | Mania ever | Psychosis ever | Depressed mood | Irritable mood | Poor concentration | Psychomotor retardation |
| Depression | | Panic_Attacks | Mania | Psychosis | m_opc37 | | m_opc41 | m_opc24 |
| Provisional_Diagnosis_5 / Provisional_Diagnosis_6 / Provisional_Diagnosis_7 / Provisional_Diagnosis_8 / Provisional_Diagnosis_9 / Provisional_Diagnosis_10 / P | | | | | | | | |
| Depression | | | Mania | Psychosis | OPCRIT1LE | | OPCRIT6LE | OPCRIT7LE |
| Depression | Anxiety | | Mania | Psychosis | cda0i01 | cda8i01 | | cdb4i01 |
| Depression | Anxiety | Panic_Attacks | | Psychosis | dep6_001 | | conc7_002 | conc7_005 |
| Depression | Anxiety | Panic_Attack | Mania | Psychosis | pda0i01 | pda8i01 | | pdb4i01 |
| Depression | Anxiety | Panic_Attacks | Mania | Psychosis | | | | |
| Depression | Anxiety | | Mania | | picdD01 | | picdD07 | picdD08 |
| Episodes_depression_LE_ever | | Panic.Disorder | Episodes_mania_ | Psychosis_LE_ | Dysphoria | | Poor_concentratior | Psychomo |
| Depression | | | | Psychosis | m_opc37 | | m_opc41 | m_opc24 |
| Depression | | | Mania | Psychosis | m_opc37 | | m_opc41 | m_opc24 |
| Depression | Anxiety | | | | | | | |
| | | | Mania | | dep_depres_mood | | dep_lack_concentr | dep_retard |
| Depression | | | Mania | | DEPR_mood | | DEPR_concentrati | DEPR_retardation |
| | | | | | S06_001 | S06_061 | S06_049 | |

**Depressi**

| Dep_Agitated_Activity | Dep_Energy | Dep_Appetite_ | Dep_Weight_Loss | Dep_Appetite_Incre | Dep_Weight_Gain |
|---|---|---|---|---|---|
| Agitated activity | Loss of energy | Loss of appetite | Weight loss | Increased appetite | Weight gain |
| m_opc23 | m_opc25 | m_opc48 | m_opc49 | m_opc50 | m_opc51 |
| Provisional_Diagnosis_11 / Provisional_Diagnosis_12 / Provisional_Diagnosis_13 / Provisional_Diagnosis_14 | | | | | |
| OPCRIT18LE | OPCRIT8LE | OPCRIT9LE | OPCRIT10LE | OPCRIT11LE | OPCRIT12LE |
| cda5i01 | cdb3i01 | cfa0i01 | cfa1i01 | cfa2i01 | cfa3i01 |
| | conc7_006 | bf8_005 - ratin | bf8_006 | bf8_005 - ratings 4 | bf8_007 |
| pda5i01 | pdb3i01 | | | | |
| | picdD03 | picdD10 | | | |
| tor_changes | Loss_of_energy | Appetite_change | | | |
| m_opc23 | m_opc25 | m_opc48 | m_opc49 | m_opc50 | m_opc51 |
| m_opc23 | m_opc25 | m_opc48 | m_opc49 | m_opc50 | m_opc51 |
| | dep_energy | appetite - one appetite variable to encompass all types of appetite disturbance | | | |
| | DEPR_energy | DEPR_appetite | | | |
| | S06_045 | S06_058 | | | |

**on**

| Dep_Ini_Insomnia | Dep_Mid_Insomnia | Dep_Waking | Dep_Sleep | Dep_Suicide | Dep_Self_Har | Dep_Interest | Dep_Self_Repr |
|---|---|---|---|---|---|---|---|
| Initial insomnia | Middle insomnia | Early morning waking | Excessive sleep | Tedium vitae / suicidal ideation (in context of depression) | History of self-harm / suicide attempts | Loss of interest or pleasure in activities | Excessive self-reproach or inappropriate guilt |
| m_opc44 | m_opc45 | m_opc46 | m_opc47 | OPCRIT_4_Suicidal_ideation | | m_opc39 | m_opc42 |
| OPCRIT13LE | OPCRIT14LE | OPCRIT15LE | OPCRIT16LE | OPCRIT4LE | | OPCRIT2LE | OPCRIT5LE |
| cfb7i02 | cfb7i03 | | cfb8i01 | cdc8i01 - scree | cdd9i01 | cdb1i01 / cdb1 | cdc3i01 |
| bf8_011 | bf8_013 | bf8_014 | bf8_016 | dep6_011 | dep6_011 | conc7_004 | dep6_013 |
| | | | | pdc8i01 | pdd9i01 | pdb1i01 /pdb1 | pdc1i01 / pdc3 |
| picdD09 | | | | picdD06 | | picdD02 | picdD05 |
| Sleep_disturbance | | | | Suicidal_ideation | | Loss_of_pleas | Excessive_self_ |
| m_opc44 | m_opc45 | m_opc46 | m_opc47 | OPCRIT_4_Suicidal_ideation | | m_opc39 | m_opc42 |
| m_opc44 | m_opc45 | m_opc46 | m_opc47 | OPCRIT_4_Suicidal_ideation | | m_opc39 | m_opc42 |
| sleep - one sleep variable to capture sleep disturbance | | | | suicide | | dep_loss_inter | dep_path_guilt |
| DEPRsleep | | | | DEPR_suicidal | | DEPR_interests | DEPR_guilt |
| S06_046 | | | S06_047 | S06_011 / S06 | S06_011 | S06_004 / S06 | S06_060 / S06_ |

| Dep_Tearfulness | Dep_Lonely | Dep_Loss_Libido | Mania_Elated_Mo | Mania_Irritable | Mania_Racing | Mania_Pressure |
|---|---|---|---|---|---|---|
| Tearfulness / crying | Loneliness | Loss of libido | Elated Mood | Irritable Mood | Racing thoughts | Over-talkativeness |
| | | m_opc40 | m_opc35 | m_opc36 | m_opc31 | m_opc30 |
| | | OPCRIT17LE | OPCRIT19LE | OPCRIT20LE | OPCRIT21LE | OPCRIT22LE |
| cda4i01 | cdb9i01 | | cde2i01 | cde4i01 | cde7i01 | cdf4i01 |
| dep6_003 | | bf8_025 | man10_001 | man10_002 | man10_004 | man10_005 |
| pda4i01 | pdb9i01 | | pde2i01 | pde4i01 | pde7i01 | pdf4i01 / pde8i01 |
| | | | pde2i01 | pde4i01 | pde7i01 | pdf4i01 / pde8i01 |
| reproach | | | Elevated_mood | Irritable_mood_ | Thoughts_racing | Pressured_speec |
| | | m_opc40 | m_opc35 | m_opc36 | m_opc31 | m_opc30 |
| | | m_opc40 | m_opc35 | m_opc36 | m_opc31 | m_opc30 |
| | | | man_elat_mood | man_irrit_mood | man_racing_th | man_overtalk |
| | | | MANI_elated | MANI_irritable | MANI_thoughts | MANI_talk |
| S06_050 | | S06_052 | | | | |

| Mania | | | | | | |
|---|---|---|---|---|---|---|
| **Mania_Distractibility** | **Mania_Overactivity** | **Mania_Esteem** | **Mania_Sleep** | **Mania_Reckles** | **Mania_Social** | **Mania_Sex** |
| Distractibility | Over-activity | Exaggerated self-esteem | Reduced need for sleep | Reckless or socially embarrassing behaviour | Increased sociability or over-familiarity | Increased sex drive |
| m_opc21 | m_opc19 | m_opc56 | m_opc22 | | m_opc53 | OPCRIT_28a_Incre |
| OPCRIT23LE | OPCRIT24LE | OPCRIT25LE | OPCRIT27LE | OPCRIT26LE | OPCRIT28LE | OPCRIT27aLE |
| cdf6i01 | cde9i01 | cdf1i01 | cdf0i01 | cdf2i01 | | |
| man10_006 | man10_007 | man10_010 | man10_013 | man10_014 | | man10_015 |
| pdf6i01 | pde9i01 / pdf5i01 | pdf1i01 | pdf0i01 | pdf2i01 | | |
| pdf6i01 | pde9i01 / pdf5i01 / pdf3i0 | pdf1i01 | pdf0i01 | pdf2i01 | | |
| Distractibility | Excessive_activity | Increased_self_esteem | Reduced_need_fo | Reckless_Activit | Increased_sociabilit | Increased_sexual_a |
| m_opc21 | m_opc19 | m_opc56 | m_opc22 | | m_opc53 | |
| m_opc21 | m_opc19 | m_opc56 | m_opc22 | | m_opc53 | |
| man_distract | mania_overact | man_self_esteem | man_reduced_sle | man_soc_emb_behav | | man_sex_activ |
| MANI_distract | MANI_overactive | MANI_esteem | MANI_sleep | MANI_embarras | | MANI_sex |

| | | | | | | | |
|---|---|---|---|---|---|---|---|
| **Hal_Aud_Abuse** | **Hal_2nd_Aud** | **Hal_3rd_Aud** | **Hal_Run_Com** | **Hal_Visual** | **Hal_Other_M** | **Thought_Insertion** | **Thought_Broadcast** |
| Abusive/ persecutory second person auditory hallucinations | Other (non-affective) second person auditory hallucinations | Third person auditory hallucinations | Running commentary | Visual hallucinations | Hallucinations in any other modality (e.g. somatic or olfactory) | Thought insertion | Thought broadcast |
| m_opc75 | m_opc76 | m_opc73 | m_opc74 | OPCRIT_33_Nona | m_opc77 | m_opc66 | m_opc68 |
| OPCRIT31 | OPCRIT32 | OPCRIT29 | OPCRIT30 | OPCRIT33 | OPCRIT34 | OPCRIT36 | OPCRIT37 |
| Psychotic symptoms (total at different waves): cpsychtot / sCpsychtot / Tcpsychtot / ppsychtot / Sppsychtot / Tppsychtot | | | | | | | |
| cjb8i01 - rating | cjb8i01 | cjb7i01 | | cjc0i01 | cjc5i01 / cjc7i0 | cjc8i01 | cjc9i01 |
| ctivity | | Third_person_AHs | | | | | Thought_interference |
| m_opc75 | m_opc76 | m_opc73 | m_opc74 | | m_opc77 | m_opc66 | m_opc68 |
| m_opc75 | m_opc76 | m_opc73 | m_opc74 | | m_opc77 | m_opc66 | m_opc68 |
| | hal_sec_third_voice | hal_sec_third_vo | hal_comment_vo | hal_vis_halluc | hal_som_hallu | thought_multi - one varia | thought_multi - one va |
| HALL_threaten | HALL_2_3_person ( | HALL_2_3_perso | HALL_comment | HALL_visual | HALL_somatic | / HALL_olfactory | |

**Psychosis**

| Thought_Withdrawal | Passivity | Del_Inf | Del_Persec | Del_Bizarre | Del_Grandios | Del_Guilt | Del_Poverty | Del_Nihilistic |
|---|---|---|---|---|---|---|---|---|
| Thought withdrawal | Passivity (replacement of will) | Delusions of influence, including reference and delusions of being spied upon. | Delusions of persecution | Bizarre delusions | Delusions of grandeur | Delusions of guilt | Delusions of poverty | Nihilistic delusions |
| m_opc67 | m_opc61 | m_opc58 | m_opc54 | m_opc59 | m_opc57 | m_opc69 | m_opc70 | m_opc71 |
| OPCRIT38 | OPCRIT39 | OPCRIT40 | OPCRIT42 | OPCRIT43 | OPCRIT55 | OPCRIT56 | OPCRIT57 | OPCRIT58 |
| | | | | | | cdc4i01 | | |
| | | | | | man10_016 (a | dep6_018 - in context of depression only | | |
| cjd1i01 | cjd3i01 | cjd4i01 | cjd5i01 | | cje1i01 | pdc4i01 / pdc4o01 / pdc4o01s | | cjd8i01 |
| | Delusions_of_passivity | | | Bizarre_delusions | | | | |
| m_opc67 | m_opc61 | m_opc58 | m_opc54 | m_opc59 | m_opc57 | m_opc69 | m_opc70 | m_opc71 |
| m_opc67 | m_opc61 | m_opc58 | m_opc54 | m_opc59 | m_opc57 | m_opc69 | m_opc70 | m_opc71 |
| thought_multi - one vari | thought_replace_will | del_reference | del_delus_pers | del_bizarre | del_grandeur | | | |
| | DELU_control | DELU_reference | DELU_persequ | DELU_bizarre | DELU_grandeu | DEPR_guilt_delusion - in context of depression o | | |
| | | | | | | S06_018 - in context of depression only | | |

| **Del_Religion** | **Del_Jealousy** | **Del_Somatic** | **Del_Hypochor** | **Del_Mind_Read** | **Del_MisID** | **Del_Other** | **Positive_FTD** | **Bizarre_Beh** |
|---|---|---|---|---|---|---|---|---|
| Religious delusions | Delusions of jealousy | Somatic delusions | Hypochondriac al delusions | Delusions of mind reading | Delusions of misidentification | Other primary delusions, including delusional mood and delusions not secondary to mood disturbance | Positive formal thought disorder | Bizarre behaviour |
| Religious_Delus | Dels_of_Jealou | Somatic_Delusi | Hypochondrical | Delusions_of_Mind_Reading_LE | | m_opc63 | m_opc28 | m_opc17 |
| OPCRIT54h | OPCRIT54g | OPCRIT54i | OPCRIT54l | OPCRIT54j | | OPCRIT44 | OPCRIT49 | OPCRIT45 |
| | | | cjd9i01 | cjd2i01 | | cjb4i01 / cje3i0 | pja3i01 | pjb0i01 |
| | | | | | | Del_Other | Positive_FTD | Bizarre_behavio |
| | | | | | | m_opc63 | m_opc28 | m_opc17 |
| | | | | | | m_opc63 | m_opc28 | m_opc17 |
| | | | dep_hypo_del (in context of depression only) | | | | formal_thought | disorgan_beh |
| nly | | | DEPR_hypochondric_delusion - in context of depression only | | | | Thought_disor | Disorganised |

| | Negative Symptoms | | | | | | | |
|---|---|---|---|---|---|---|---|---|
| **Psychotic_Exp** | **Negative_FTD** | **Flat_Affect** | **Blunt_Affect** | **Inapp_Affect** | **Poor_Grooming** | **Impersistence** | **Few_Recreation** | **Intimacy** |
| Experiences of psychotic experiences / prodromal psychosis | Negative formal thought disorder | Restricted affect | Blunted affect | Inappropriate affect | Poor grooming and hygiene | Impersistence at work or school | Few or no recreational interests and activities | Inability to form close or intimate relationships (romantic or family) |
| | m_opc29 | m_opc32 | m_opc33 | m_opc34 | OPCRIT_54b_Po | OPCRIT_54c_Impe | OPCRIT_54d_Recre | OPCRIT_54e_Abilit |
| | | OPCRIT51 | OPCRIT52 | OPCRIT53 | OPCRIT54b | OPCRIT54c | OPCRIT54d | OPCRIT54e |
| | | cda9i01 | | | | | | |
| Psychotic_Experiences | | pda9i01 | | | | | | |
| ur | | | | | | | | |
| | m_opc29 | m_opc32 | m_opc33 | m_opc34 | | | | |
| | m_opc29 | m_opc32 | m_opc33 | m_opc34 | | | | |
| | NEGA_speech | NEGA_affective - | NEGA_affective - covers restri | | NEGA_appearan | Work | Lack_interests | |
| | S06_057 - in con | S06_008 - in context of depression only | | | | | | |

| Friends | ADHD_Atten | ADHD_Mistakes | ADHD_Forget | ADHD_Persisten | ADHD_Instruct | ADHD_Tasks | ADHD_Distrac |
|---|---|---|---|---|---|---|---|
| Few or no friends | Short attention span | Careless Mistakes | Forgetfulness / losing things | Lack of persistence | Difficulty listening to or carrying out instructions | Changing tasks or activities | High distractibility |
| OPCRIT_54f_Relationships_with_friends__peers__LE | | | | | | | |
| OPCRIT54f | | | | | | | |
| | prb4i01 | prc6i01 | prc8i01 / prb2i01 | prc5i01 | pra8i01 / prb3i01 | | pra9i01 |
| | prb4i01 | prc6i01 | prc8i01 / prb2i01 | prc5i01 | pra8i01 / prb3i01 | | pra9i01 |
| | ADHD_IN01ev | ADHD_IN07ever | ADHD_IN05ever / A | ADHD_IN08ever | ADHD_IN02ever / ADH | ADHD_IN09ever | ADHD_IN04eve |
| social_isolation? | | | | | | | |

| ADHD | | | | | | | |
|---|---|---|---|---|---|---|---|
| **ADHD_Sitting** | **ADHD_Fidget** | **ADHD_Concen** | **ADHD_Activity** | **ADHD_Speech** | **ADHD_Turn** | **ADHD_Thinking** | **ADHD_Interrupt** |
| Inability to sit still | Frequent fidgeting | Poor concentration | Excessive physical movement / activity | Excessive speech | Inability to wait their turn | Acting without thinking | Interrupting conversations |
| pra2i01 | pra0i01 | pra7i01 | prc4i01 | pra5i01 | prb7i01 | | prc1i01 |
| pra2i01 | pra0i01 | pra7i01 | prc4i01 | pra5i01 | prb7i01 | | prc1i01 |
| ADHD_H02ever | ADHD_H01ever | | ADHD_H03ever / ADHD_I | ADHD_IM04ever | ADHD_IM01ever | | ADHD_IM02ever / |

| | | | | | | | Anxiety |
|---|---|---|---|---|---|---|---|
| **Anx_Affect** | **Anx_Tension** | **Anx_Restless** | **Anx_Breath** | **Anx_Heart** | **Anx_GI** | **Anx_Vigilance** | **Anx_Sleep** |
| Anxious affect | Muscle tension | Restlessness | Breathing difficulties / hyperventilation | Palpitations / tachycardia | Gastrointestinal problems | Hypervigilance | Sleep disturbance due to anxiety |
| Anxiety symptoms (total at different waves): canxGAD / ScanxGAD / TcanxGAD / panxGAD / SpanxGAD / TpanxGAD | | | | | | | |
| pca3i01 / pca4i01 | pcd0i14 | pcd0i21 | pce5i04 / pce5i05 | pce5i06 | pce5i18 | pcc2i01 / pcd0i20 | |
| | | | | | | Anxiety symptoms available - specific to childho | |

| | | | | | | | | |
|---|---|---|---|---|---|---|---|---|
| | | | | | | **Oppositional I** | | |
| **Anx_Concentrati** | **Anx_Avoid** | **Anx_Agora** | **Anx_Phobia** | **Panic_Attacks** | **Anx_Sep** | **Anger** | **Defiance** | **Vindictive** |
| Poor concentration due to anxiety | Avoidance behaviours | Agoraphobia | Specific phobias | Panic attacks | Separation anxiety | Angry / irritable mood | Argumentative / defiant behaviour | Vindictiveness |
| | | | | Panic_age_1st_impairment | | | | |
| pcc3i01 | Individual variables for | pcb2i01 | pca5i01 | pcc5i01 | pbe7i01 | pge0i01 | pga1i01 | pga3i01 |
| ood | | | | | | ODD_01p / OD | ODD_02p / OD | ODD_04p / ODD_( |

| Defiant / Conduct Disorders | | | | Autism | | | | |
|---|---|---|---|---|---|---|---|---|
| **Aggression** | **Destroy** | **Deceit** | **Rule_Violation** | **Socio_Emo** | **Verbal_Comn** | **Nonverbal_Com** | **Relationships** | **Rep_Move** |
| Aggression towards people / animal | Destruction of property | Deceitfulness / theft | Serious violations of rules | Deficits in socio-emotional reciprocity | Deficits in verbal communication | Deficits in nonverbal communication | Deficits in developing and maintaining relationships | Stereotyped or repetitive movements |
| | | | | AQ_Total | | | | |
| pge5i01 / pge6i01 | pge1i01 | pga5e01 / pga | pga0i01 / pga1x0 | ASQ variables - do not map exactly onto thes | | | | |
| CD_10p / CD_11 | CD_15p / CD_ | ODD_05p / CD | CD_12p / CD_18 | Autism Quotient | | | | |

| | | | **Irritability** | | | | | |
|---|---|---|---|---|---|---|---|---|
| **Inflexibility** | **Fixed_Interes** | **Sensory_Reactivity** | | **Emo_SDQ3** | **Emo_SDQ8** | **Emo_SDQ13** | **Emo_SDQ16** | **Emo_SDQ24** |
| Inflexible adherence to routines / ritualised patterns of behaviour | Highly restricted, fixated interests | Hyper- or hypo-reactivity to sensory input or unusual interest in sensory aspects of the environment | Measure of irritability not within the context of depression or mania - mainly for childhood studies | Complains of headaches (0/1/2) | Often worries (0/1/2) | Often unhappy (0/1/2) | Nervous or clingy (0/1/2) | Many fears (0/1/2) |
| | | | | | | SDQ_Emotional | | |
| | | | TEMPS: ctemps21, ctemps22, ctemps23, ctemps24, ctemps25, ctemps26, ctemps27, ctemps28 | | | | | |
| | | | TEMPS: mtemps21, mtemps22, mtemps23, mtemps24, mtemps25, mtemps26, mtemps27, mtemps28, ftemp | | | | | |
| se | | | | P_SDQ_3 | P_SDQ_8 | P_SDQ_14 | P_SDQ_17 | P_SDQ_26 |
| | | | Covered by depression | | | | | |

**Strengths and Difficulties Que**

| Conduct_SDQ | Conduct_SDQ7 | Conduct_SDQ12 | Conduct_SDQ18 | Conduct_SDQ22 | Hyper_SDQ2 | Hyper_SDQ10 | Hyper_SDQ15 |
|---|---|---|---|---|---|---|---|
| Anger / temper tantrums (0/1/2) | Generally obedient (2/1/0) | Often fights (0/1/2) | Often lies or cheats (0/1/2) | Often steals (0/1/2) | Restlessness, overactivity (0/1/2) | Constant fidgeting or squirming (0/1/2) | Easily distracted (0/1/2) |
| SDQ_Conduct | | | | | SDQ_Hyper | | |
| ps21, ftemps22, ftemps23, ftemps24, ftemps25, ftemps26, ftemps27, ftemps28 | | | | | | | |
| P_SDQ_5 | P_SDQ_7 | P_SDQ_12 | P_SDQ_19 / P_SD | P_SDQ_23 / P_SD | P_SDQ_2 | P_SDQ_10 | P_SDQ_16 |
| | | | | | Not available at baseline - when was this dor | | |

**estionnaire**

| Hyper_SDQ21 | Hyper_SDQ25 | Peer_SDQ6 | Peer_SDQ11 | Peer_SDQ14 | Peer_SDQ19 | Peer_SDQ23 | Prosocial_SD | Prosocial_SDQ4 |
|---|---|---|---|---|---|---|---|---|
| Thinks things out before acting (2/1/0) | Sees tasks through to the end (2/1/0) | Solitary, tends to play alone (0/1/2) | At least one good friend (2/1/0) | Generally liked by other children (2/1/0) | Bullied (0/1/2) | Gets on better with adults than children (0/1/2) | Considerate of other people's feelings (0/1/2) | Shares readily with other children (0/1/2) |
| | | SDQ_Peers | | | | | | |
| P_SDQ_30 | P_SDQ_31 | P_SDQ_6 | P_SDQ_11 | P_SDQ_15 | P_SDQ_21 | P_SDQ_25 | P_SDQ_1 | P_SDQ_4 |
| ie and can we request? | | | | | | | | |

| Prosocial_SDQ9 | Prosocial_SDQ17 | Prosocial_SDQ20 | AD | Anxio | Hyp | MoodStabil | AP |
|---|---|---|---|---|---|---|---|
| Helpful if someone is hurt (0/1/2) | Kind to younger children (0/1/2) | Often volunteers to help others (0/1/2) | Anti-depressants ever | Anxiolytics ever | Hypnotics ever | Mood stabilisers ever | Antipsychotics (oral) ever |
| | | | Antidepressants_Eve | Anxiolytics_eve | Hypnotics_eve | Mood_Stabilise | Antipsychotics |
| SDQ_Prosocial | | | Antidepressants | Anxiolytics | Hypnotics | Mood_stabilise | Antipsychotics |
| | | | Antidepressants | Anxiolytics | Hypnotics | Mood Stabilise | Antipsychotics |
| P_SDQ_9 | P_SDQ_18 | P_SDQ_22 | MED_1 / MED_2 / MED_3 / MED_4 | | | | |
| | | | Description_of_histor | | | | |
| | | | med_assess | | | | |
| | | | Antidepressants_eve | Anxiolytics_ev | Hypnotics_eve | Mood_stabilise | AP_e |
| | | | m_meds_antidep_cu | m_meds_anx_ | current - no life | time variable | m_meds_antipsych_ |
| | | | m_meds_antidep_cu | m_meds_anx_ | current - no life | time variable | m_meds_antipsych_ |
| | | | | | | | neuro_treat |
| | | | | | | | AP |
| | | | AD | | | | |

| History of Treatment | | | | | | | | |
|---|---|---|---|---|---|---|---|---|
| **Depot** | **ECT** | **CBT** | **Other_Talkin** | **Lithium** | **Clozapine** | **ADHD_Med** | **Treatment_Re** | **Age_Onset** |
| Antipsychotics (depot) ever | ECT ever | CBT ever | Other talking therapy than CBT ever | Lithium ever | Clozapine ever | ADHD medication ever (stimulant or nonstimulant) | Variables relating to treatment response | |
| Depots | ECT | CBT | Other_Talking | Lithium_Respons | Clozapine_Ever | | m_opc89 | Age_Onset |
| | ECT | CBT | Other_talking_ | Lithium | Clozapine | ADHD_stimulants | | age_of_onset / age_of_im |
| | ECT | CBT | Other_talking_ | Lithium | Clozapine | ADHD_stimula | OPCRIT89 | age_of_onset / age_of_im |
| | | | | | | | | Age_Onset |
| / MED_5 / MED_6 / MED_7 / MED_8 / MED_9 / MED_10 / MED_11 / MED_12 | | | | | | | | |
| ic_medication / Current_medication_description | | | | | | | | Age_at_onset_of_psychot |
| s / med_assess_details | | | | | | med_stim / risperidone | | ADHD_onset |
| ver | ECT_ever | CBT_ever | Other_talking_ | Lithium_ever_tak | Clozapine_ever_taken | | | Age_onset_impairment |
| current - no lifetime variable | | | | | | | m_opc89 | Age_Onset |
| current - no lifetime variable | | | | | | | m_opc89 | Age_Onset |
| | | CBT | Other_Talking | | | | | |
| | | | | li_treat | | | | ill_onset_imp |
| | ECT | | | Lithium | | | Neuroleptic_res | Onset_impairment |
| | | | | | | | S06_034 | Age_at_onset_for_MDD / |

**Clinical Variables**

| Age_Treatment | Duration_Illness | Duration_Treatm | Admissions | Longest_Adm | Ever_Sectioned | Age_1st_Admiss |
|---|---|---|---|---|---|---|
| | In weeks | | Number of hospital admissions | Longest admission in weeks | Ever sectioned | Age of first hospital admission |
| Age_Treatment | m_opc8 | antipsychotic_expo | Number_of_admissions | | Ever_Sectioned | Age_Admission |
| age_first_treatment | | | number_of_admissions | longest_admission_weeks | | |
| age_first_treatment | | | number_of_admissions | longest_admission_weeks | | |
| | | | Mhospital / Fhospital | | | |
| ic_disorder | | | | | | |
| Age_Treatment_Stim / Age_Treatment_Other | | | | | | |
| | | | Number_of_admissions | | Ever_sectioned | Age_onset_admiss |
| | m_opc8 | | Number_of_admissions | | Ever_Sectioned | Age_Admission |
| | m_opc8 | | | | | |
| | CAPSduration_months | | | | | |
| first_psy | | | num_hosp | | | first_hosp |
| | | Years_neuroleptics | Hospitalisations_N | | | Onset_Hospitalisat |
| S06_025_1 | S06_026d_1 - duration of representative episode of depression | | | | | |

| Eps_Psychosis | Eps_Dep | Eps_Mania | Course_Illness | Mode_Onset | Sleep | Suicidal_Ideat |
|---|---|---|---|---|---|---|
| Number of episodes of psychosis | Number of episodes of depression | Number of episodes of mania | Course of illness | Mode of onset | Sleep disturbance independent of illness | History of suicidal ideation / attempts |
| Total_Psychotic_Episo | Number_of_episodes_depre | Number_of_episodes | m_coursedisorder | m_mode_onset | | Suicidal_ideati |
| Num_Episodes_Psycho | Number_of_Depression_Ep | Num_Episodes_High | OPCRIT90 | OPCRIT5 | | OPCRIT4LE |
| | | | | | Sleep section o | cdc8i01 |
| | | | mcourse / fcourse - cour | se of depression | | dep6_011 - on c |
| | | | | | Sleep section o | Suicide section |
| | | | | | Sleep section o | Suicide section |
| sion | Number_of_episodes_depre | Number_of_episodes | _mania_LE | | | Suicidal_Ideati |
| | | | m_coursedisorder | m_mode_onset | | |
| | | | m_coursedisorder | m_mode_onset | | |
| | | | | | Pittsburgh1 / Pit | BPD5 |
| | | man_num_episodes | quality_remissions | | | suicide - only in |
| ion | | MANI_N_episodes | Remission_type | Onset_type | | DEPR_suicidal |
| | S06_030_1? | | S06_033_1 | | | S06_011 - only |

| | | | | | | | |
|---|---|---|---|---|---|---|---|
| **Death_Parent** | **Death_Parent_** | **Death_Sibling** | **Death_Sibling_** | **Death_Friend** | **Death_Friend_** | **Parent_Separa** | **Parent_Separate_Age** |
| CLEQ_item_1_yes_( | CLEQ_item_1_y( | CLEQ_item_2_y( | CLEQ_item_2_y( | CLEQ_item_3_y( | CLEQ_item_3_y | CLEQ_item_4_y | CLEQ_item_4_years (di |
| | | | | | | ace_calculator_7 | |
| | | | | | | ace_calculator_7 | |
| ontext of depression only | | | | | | | |
| P_Life_Events_8 | | P_Life_Events_8 | | P_Life_Events_10 | | P_Life_Events_6 | |
| of CAPA - suicide screening question: pdc8i01 | | | | | | | |
| on_LE | | | | | | | |
| ACE15 | | | | | | ACE6 | |
| context of depression | | | | | | | |
| Bleq_03 / Bleq_04 - Death of loved ones | | | | | | | |

| Serious_Illness | Serious_Illness_ | Parent_Illness | Parent_Illness_ | Visible_Deform | Visible_Deformity_ | Parent_Prison | Parent_Prison_ |
|---|---|---|---|---|---|---|---|
| CLEQ_item_7_yes_no | CLEQ_item_7_ye | CLEQ_item_8_ye | CLEQ_item_8_y | CLEQ_item_9_y | CLEQ_item_9_years | CLEQ_item_10_yes | CLEQ_item_10_ |
| lec_12 | | | | | | ace_calculator_10 | |
| lec_12 | | | | | | ace_calculator_10 | |
| P_Life_Events_24 | | P_Life_Events_5 | | | | P_Life_Events_18 | |
| LEC14 | | | | | | ACE10 | |
| Bleq_01 | | Bleq_02 - Serious | illness of a loved | one | | | |

**Life Events**

| Teenage_Paren | Teenage_Parent_ | School_Suspensio | School_Suspension_ | Physical_Ab | Physical_Ab_ | Sexual_Ab | Sexual_Ab_A | Emotional_Al |
|---|---|---|---|---|---|---|---|---|
| CLEQ_item_11_ | CLEQ_item_11_ye | CLEQ_item_12_ye | CLEQ_item_12_years | CLEQ_item_13_yes_ | CLEQ_item_1 | CLEQ_item_1 | CLEQ_item_13_years | |
| | | | | ace_calculator_2 | | ace_calculator_3 | | ace_calculato |
| | | | | ace_calculator_2 | | ace_calculator_3 | | ace_calculato |
| | | | | | | | | P_Life_Events_ |
| | | | | ACE2 | | ACE3 | | ACE1 |

| **Emotional_Ab_** | **Neglect** | **Neglect_Age** | **Witness_Viol** | **Witness_Violence_** | **Parent_Substance** | **Parent_Substance_** | **Serious_Accident** |
|---|---|---|---|---|---|---|---|
| r_1 | ace_calculator_ | 5 | ace_calculato | r_6 | ace_calculator_8 | | lec_4 |
| r_1 | ace_calculator_ | 5 | ace_calculato | r_6 | ace_calculator_8 | | lec_4 |
| _23 - More criticis | P_Life_Events_2 | 2 | | | | | |
| | ACE5 | | ACE7 | | ACE8 | | LEC4 |

| | | | | | | | | | |
|---|---|---|---|---|---|---|---|---|---|
| | | | | | | | **Education &** | | |
| **Serious_Accident_** | **Disaster** | **Disaster_Age** | **Fire_Explo** | **Fire_Explo_A** | **Witness_Dea** | **Witness_Dea** | **Pt_Edu** | **Mother_Edu** | **Father_Edu** |
| | | | | | | | m_edu_attainm | Mother_Highes | Father_Highe |
| | lec_1 | | lec_2 | | lec_14 | | Education_Attaintment | | |
| | lec_1 | | lec_2 | | lec_14 | | Education_Attaintment | | |
| | | | | | | | | meducationGC | feducationGCS |
| | | | | | | | meducationGCSE / meducationAlevel / medu | | |
| | | | | | | | Pt_Edu | | |
| | | | | | | | Pt_Edu | Mother_Edu | |
| | | | | | | | | Education | |
| | | | | | | | Highest_educational_attainment | | |
| | | | | | | | m_edu_attainment | | |
| | | | | | | | m_edu_attainment | | |
| | LEC1 | | LEC2 | | LEC16 | | Education | | |
| | | | | | | | cont_edu | | |
| | | | | | | | Higher_education - only codes whether they v | | |

| Occupation | | | | | | | | | |
|---|---|---|---|---|---|---|---|---|---|
| **Pt_Occ** | **Mother_Occ** | **Father_Occ** | **Smoking** | **Age_Smoking** | **Heaviest_Sm** | **Alcohol_Heaviest** | **Alc_Problems** | **Alc_Ab_Dep** | **Cannabis** |
| | | | Ever regular smoker | | Number of cigarettes at heaviest | Heaviest units per week | Problems at heaviest | | Cannabis ever regular |
| Highest_Occup | m_motheroccu | m_fatheroccup | Ever_regular_ | Age_first_started_to_smoke | | m_alcohol_units_h | m_alc_heavies | m_opc78 | Cannabis |
| | | | Ever_regular_ | Age_began_to | Ave_cigarettes | Alcohol_units_per_week_at_heav | | Alcohol_harmf | Cannabis_ever |
| | | | Ever_regular_ | Age_began_to | Ave_cigarettes | Alcohol_units_per_week_at_heav | | Alcohol_harmf | Cannabis_ever |
| E / feducationAlevel / feducationdegree / feducationhigher / feducationvocation | | | | | | | | | |
| cationdegree / meducationhigher / meducationvocation / feducationGCSE / feducationAlevel / feducationdegree / feducationh | | | | | | | | Alc_Ab_Dep | |
| | P_Edu_5 | P_Edu_6 | Smoking | Age_Smoking | pha0v01 | pha3i01 - not heaviest (CAPA only | | severityalc? | phboe01 |
| Employment | | | | | | | | Alcohol_use_disorder | |
| | Social_status_FINAL | | smoking3 | | smoking4 - ma | alcohol2 / alcohol3 | | | cannabis2 - tim |
| Highest_Occupation | | | Ever_regular_smoker | | | Heaviest_alcohol_units_per_wk | | | Cannabinoids_ |
| | m_motheroccu | m_fatheroccupation | | | | m_alcohol_units_h | m_alc_heavies | m_opc78 | Cannabis |
| | m_motheroccu | m_fatheroccupation | | | | m_alcohol_units_h | m_alc_heavies | m_opc78 | Cannabis |
| Currently_employed / Current_job - no lifetime variables | | | | | | Could derive a rou | Audit3 / Audit4 | Alc_Ab_Dep | Drugs_year_ca |
| | | | | | | | | alco_intake | |
| went on to higher education | | | | | | | | Alcohol | |
| | | | | | | | | Alcohol_dependency_disorde | |

**Substance Use**

| Can_Problem | Can_Ab_Dep | Other_Drug | OD_Problems | OD_Ab_Dep | Opioids | Amphetamine | Sedatives | Hallucinogens |
|---|---|---|---|---|---|---|---|---|
| | | | | | Opioids ever regular | Amphetamines ever regular | Sedatives ever regular | Hallucinogens ever regular |
| Cannabinoids_ | m_opc79 | Unspec_drugs_ | Unspec_drug_p | m_opc80 | Opioids | Amphetamines | Sedatives | Hallucinogens |
| _regular | Cannabis_harr | Other_drug_use_ever_regular | | Other_drug_harmful_use_or_dependence | | | | |
| _regular | Cannabis_harr | Other_drug_use_ever_regular | | Other_drug_harmful_use_or_dependence | | | | |
| | | Not derived for this dataset as: | | severitydrug? | phb6e01 / phb7 | phb3e01 | phc1e01 | phb8e01 / phc0 |
| | Cannabis_use_disorder | | | OD_Ab_Dep | | | | |
| es taken - could be used to de | | drugsother1 - not regular | | | | Amphetamines | | Hallucinogens |
| ever_regular | | Unspec_drugs_ever_regular | | | | | | |
| SPq104 | m_opc79 | | SPq99 / SPq10 | m_opc80 | Opioids | Amphetamines | Sedatives | Hallucinogens |
| | m_opc79 | | | m_opc80 | Opioids | Amphetamines | Sedatives | Hallucinogens |
| Fq_use_cannabis - not exactly | | Past year only | Past year only: Fq_use_amphetamines / Fq_use_cocaine / Fq_use_crack / Fq_use_ecstacy / Fq_use | | | | | |
| | | | | drug_ab | | | | |
| | | | | Drugs | | | | |

| | | **Family History** | | | | | | | |
|---|---|---|---|---|---|---|---|---|---|
| **Cocaine** | **Ecstasy** | **FH_Psychosis** | **FH_Schizophre** | **FH_Bipolar** | **FH_Dep** | **FH_Suicide** | **FH_ID** | **FH_ADHD** | **FH_Genetic** |
| Cocaine ever regular | Ecstasy ever regular | 1st/2nd degree family history of psychosis | 1st/2nd degree family history of schizophrenia | 1st/2nd degree family history of bipolar disorder | 1st/2nd degree family history of unipolar depression | 1st/2nd degree family history of suicide | 1st/2nd degree family history of intellectual disability | 1st/2nd degree family history of ADHD | 1st/2nd degree family history of genetic syndrome |
| Cocaine | Ecstasy | FHPsychosis_ | OPCRIT D&H 13 | FHBipolar_1st_or_2 | FHDepression_ | FHsuicide_1st_ | FH_Other_LD | | |
| | | family_psychosis_schizophrenia | | family_bipolar | family_depressi | family_suicide | family_LD | family_ADHD | family_genetic |
| | | family_psychosis_schizophrenia | | family_bipolar | family_depressi | family_suicide | family_LD | family_ADHD | family_genetic |
| | | FH_Psychosis | | | FH_Dep | fhsuicide | | | |
| phb1e01 / phb2e01 | | | | | | | | | |
| | | FH_Psychosis | FH_Schizophren | FH_Bipolar | FH_Dep | | FH_ID | FH_ADHD | |
| Cocaine | Ecstasy | Family history section - specifies whether specific family members have a mental health problem - specific disorders are free tex | | | | | | | |
| Cocaine | | | m_opc13 | | | | | | |
| Cocaine | | | | | | | | | |
| _heroin / Fq_use_LSD / Fq_use_mushrooms / Fq_use_methadone / Fq_use_semeron / Fq_use_benzos / Fq_use_amyl / Fq_use_steroids / Fq_use_solvents / Fq_ | | | | | | | | | |
| | | fam_hist - string variable | | | | | | | |
| | | FH_Psychosis | FH_Schizophren | FH_Bipolar | FH_Dep | | | | |

| FH_Autism | HD_Ischemic | HD_Congenital | Diabetes_I | Diabetes_II | Hypertension | High_lipids | Seizures | Migraines | Head_injury |
|---|---|---|---|---|---|---|---|---|---|
| 1st/2nd degree family history of autism | Ischemic heart disease | Congenital heart disease | Diabetes - type I | Diabetes - type II | High blood pressure | High cholesterol | Seizures | Migraines | Head injury |
| FH_Autism | Heart_Disease | | m_diabetes - ratin | m_diabetes - ra | Hypertension | high_cholester | epilepsy | Migraine | m_headtrauma |
| family_autism | Heart_Disease | | Diabetes_Type_1 | Diabetes_Type | Hypertension_ | Elevated_lipids_ | Epilepsy_Seizu | Migraine_Head | Head_Injury |
| family_autism | Heart_Disease | | Diabetes_Type_1 | Diabetes_Type | Hypertension_ | Elevated_lipids_ | Epilepsy_Seizu | Migraine_Head | Head_Injury |
| | | P_Health_dev_24 | | | | | P_Health_dev_ | 16 | P_Health_dev_ |
| FH_Autism | Cardiac_problems_ | | Diabetes - all types | | Description_of_ | Description_of_c | Unprovoked_seizures_ | | To cover all ne |
| xt variables | | | | | | | DH_epilepsy | | DH_headinjury |
| | Heart_Disease | | Diabetes_Type_I_or_II | | Hypertension | High_lipids | Seizures | Migraine_Headaches | |
| | | | m_diabetes - ratin | m_diabetes - rating 2 | | | c_neurological_y - rating 3 | | m_headtrauma |
| | | | m_diabetes - ratin | m_diabetes - rating 2 | | | c_neurological_y - rating 3 | | m_headtrauma |
| use_ketamine / Fq_use_legal / Fq_use_other | | | | | | | Nonepileptic_s | Migraine | |
| | | | Diabetes - all types | | | | Epilepsy / fits | | Head_injury |
| | Heart_attack_or_angina | | Diabetes_type_1 | Diabetes_type | Hypertension | High_cholestero | Epilepsy_or_co | Migraine_headaches | |

**Physical Health**

| **Stroke** | **Thyroid** | **Cancer** | **Kidney_Prob** | **Liver_Prob** | **Dementia** | **Osteoarthritis** | **Osteoporosis** | **Meningitis** | **Asthma** |
|---|---|---|---|---|---|---|---|---|---|
| Stroke or haemorrhage | Thyroid disease | Cancer | Kidney problems | Liver problems | Dementia | Osteoarthritis | Osteoporosis | Meningitis / encephalitis | Asthma |
| Stroke | Thyroid_Disease | Cancer | kidney_probl | liver_problems | Dementia | | | Meningitis_Enc | Asthma |
| Stroke_Haemo | Overactive_thyroi | Breast_cancer | Kidney_Diseas | Liver_Disease | Memory_Loss_ | Osteoarthritis | Osteoporosis | Meningitis | Asthma |
| Stroke_Haemo | Overactive_thyroi | Breast_cancer | Kidney_Diseas | Liver_Disease | Memory_Loss_ | Osteoarthritis | Osteoporosis | Meningitis | Asthma |
| _14 | P_Health_dev_26 (thyroid problems) | | | | | | | | |
| urological: Any | Description_of_endocrine_disord | | Genitourinary_problems | | | Osteoarthritis | Osteoporosis | | |
| | | | | | | | | | DH_asthma |
| Stroke | Thyroid_Disease | Cancer | Kidney_Diseas | Liver_Disease | Memory_Loss_ | Osteoarthritis | Osteoporosis | | Asthma |
| SPq157 | c_hormonal | | | | | | | | SPq151 - ratin |
| [illegible] | c_hormonal | | | | | | | | |
| | Thyroid | | | | | | | | |
| Stroke | Thyroid_disease | Cancer | Kidney_diseas | Liver_disease | Dementia_or_r | Osteoarthritis | Osteoporosis | | Asthma |

| | | | | | | | |
|---|---|---|---|---|---|---|---|
| | | | **Functioning** | | | | |
| **MS** | **Parkinson** | **Rheum_arth** | **GAS_Worst** | **GAS_Intervie** | **Current_Work** | **Ever_Married** | **Income** |
| Multiple sclerosis | Parkinson's disease | Rheumatoid arthritis | Global Assessment Scale Score - Worst episode | Global Assessment Scale Score - At Interview | Current occupation - (working, not working due to sickness, unemployed | Relationship status at time of interview | |
| MS | Parkinsons_D | Rheumetoid_ | GAS_Worst | GASPastWeek | Current_Occupat | m_marital_history | |
| Multiple_Sclero | Parkinson_Dis | Rheumatoid_Arthritis | | | Employment | ever_married | GrossIncome |
| Multiple_Sclero | Parkinson_Dis | Rheumatoid_A | GAS_Worst | GAS_PAST_W | Employment | ever_married | GrossIncome |
| | | | | | | | famincome1 / famincom |
| | | | | | | | famincome1 / famincom |
| | | | | | | | P_Edu_2 |
| | | Rheumatoid_arthritis | | GAF | Current_Work | | |
| | | | | | | | Income |
| Multiple_Sclero | Parkinson_Dis | Rheumatoid_A | GAS_Worst | | Current_Work | | |
| c_neurological_ | c_neurological_y - rating 2 | | | | | m_marital_history | |
| c_neurological_ | c_neurological_y - rating 2 | | | | | m_marital_history | |
| | | | CAPS_Impair_Occ_Fn / CAPS | | Currently_employe | Marital_status | |
| | | | | | prof | mar_status | |
| | | RA | | | Profession / Social | Married | |
| Multiple_sclero | Parkinsons_dis | Rheumatoid_arthritis | | | | | |

| | | Cognition |
|---|---|---|
| **Children** | **Deterioration_Premo** | **Cognitive_g** |
| | Deteriorated from premorbid level of functioning | General Intelligence 'g' |
| Number_of_children | m_opc88 | MCCB variables |
| num_of_children | | Online cog variables |
| num_of_children | | Online cog variables |
| e2 / famincome3 | | cIQ / cwssvc / cwsswm / cwssps / cwsspr |
| e2 / famincome3 | | |
| | | CANTAB and other child/adult tasks |
| | | VIQ / PIQ / FSIQ |
| | | WISC - CANTAB also available |
| | m_opc88 | |
| | m_opc88 | |
| Children | | |
| Children | | |
